# Project Prevent: Assessing the Value of Education-Focused Laws in Reducing Child Sexual Abuse

**DOI:** 10.1101/2022.01.31.22270166

**Authors:** Bradley Merrill Thompson, Kunal Srivastava

**Author notes:** **Appendix A—Detailed Analysis** The authors are graduate students at the University of Michigan School of Information (“UMSI”) pursuing a Master of Applied Data Science, and did this research as their capstone project. In our project, we were joined by Lindsay Durand, MPH, MADS. Ms. Durand took the lead in conducting our social media analysis where we tried to find evidence of the prevalence of educational programs addressing child sexual abuse. She also gathered and analyzed demographic data for the project, and created many of the visualizations for the raw data. We are extremely grateful and felt privileged to work alongside Ms. Durand on the project. The authors would like to thank Professor Scott Cunningham at Baylor University for his insights into causal inference. At UMSI, several of the teaching staff helped nurture the original idea for the study and were sounding boards for conceptualizing and implementing it. They include, in a course on Causal Inference, Alain Cohn, in Social Media Analysis, Paul Resnick and in Data Science for Social Good, Yan Chen, Linfeng Li, and Zhewei Song. We would also like to thank a former UMSI graduate student, now an Assistant Professor at the University of Minnesota, Carlson School of Business, Teng Ye, for guiding us on the proper use of a causal random forest. Last but not least, we would like to thank our UMSI faculty advisors, Qiaozhu Mei and Elle O’Brien, for helping us navigate the University process for contracting for the data and the institutional review board review. The contracts for the study were managed by the University of Michigan Office of Research and Sponsored Projects, under an Unfunded Agreement (“UFA”) (22-UFA0597). The National Child Abuse and Neglect Data System (NCANDS) is a voluntary data collection system that gathers information from all 50 states, the District of Columbia, and Puerto Rico about reports of child abuse and neglect. NCANDS was established by the Children’s Bureau of the US Department of Health and Human Services in response to the Child Abuse Prevention and Treatment Act of 1988. The analyses presented in this publication are based on data from the 2005-2019 NCANDS Child File. The National Data Archive on Child Abuse and Neglect (“NDACAN”) at Cornell University provided the data, and gave permission for the study. The University of Michigan acquired access to the data through a license from NDACAN and the data are subject to certain restrictions. The collector of the original data, NDACAN, Cornell University, and the agents or employees of these institutions bear no responsibility for the analyses or interpretations presented here. The information and opinions expressed reflect solely the opinions of the authors. This study is being conducted under the oversight of the University of Michigan Health Sciences and Behavioral Sciences Institutional Review Board (“IRB-HSBS”): Project Prevent: Assessing the Value of Education-Based Legislative Interventions Aimed At Reducing Child Sex Abuse Study eResearch ID: HUM00204536 Initial IRB Approval Date: 9/3/2021 Our code used in the study is available on a GitHub repository at: https://github.com/BradleyMerrillT/ProjectPrevent.

## Abstract

We investigated the impact of new state laws requiring or encouraging education on Child Sexual Abuse (“CSA”) in schools. From 2005 through June, 2019, 31 states enacted such laws. By comparing the states with and without those laws using a difference-in-differences calculation, and controlling for changes in population, wages, employment and other relevant CSA laws, we conclude that most likely such laws:

- *Increased* the number of CSA reports made to state departments of child services each year, and
- *Decreased* the number of CSA cases that those departments were able to substantiate from the reported cases.

We had hypothesized, however, that effective educational content would cause more reported cases to be substantiated as people learn what to report, but implementation of the new laws apparently instead increased the proportion of *unsubstantiated* reports of CSA.

Our research does not tell us whether that is good or bad. In the best scenario, children are helped, but at the expense of potentially hurting those who are wrongly accused. In another scenario, children and the wrongly accused are hurt. In arguably the worst scenario, child are hurt and perpetrators are helped.

We recommend that policymakers support research to learn why the laws are shifting more cases to unsubstantiated. Depending on the answer, the laws may be worthwhile to continue but with a modification to how the reporting is done, creating a new path for reporting troublesome but lawful activity. Such a modification would preserve the benefit to children while avoiding the harm to adults.

## II. Introduction

Our goal is to assess the impact of certain educational laws on the rate of child sexual abuse (“CSA”). In particular, we wish to respond to the observation of the Centers for Disease Control and Prevention (“CDC”) that “[r]esources for CSA have mostly focused on treatment for victims and criminal justice-oriented approaches for perpetrators. While these efforts are important after CSA has occurred, little investment has been made in primary prevention, or preventing CSA before it occurs. Limited effective evidence-based strategies for proactively protecting children from CSA are available.”^1^

Since about 2009, various advocates including both nonprofits and survivors of CSA have been urging state legislatures across the country to adopt laws designed to prevent CSA. Through 2019, 31 state legislatures have adopted laws that focus on educating children and sometimes staff and parents on how to identify CSA and report it. Throughout this article, we refer to these as the CSA educational laws.

Making our analysis of those laws possible, just this past June, as a part of the Enough Abuse Campaign, the Massachusetts Citizens for Children conducted a comprehensive review of state laws that seek to prevent CSA and grouped them into categories. They published their report, entitled “A Call to Action for Policymakers and Advocates: Child Sexual Abuse Prevention Legislation in the States,” in June 2021.^2^ Our plan is to quantitatively assess the impact of the CSA educational laws identified in that report.

## III. CSA Deeply Hurts Children

The CDC has synthesized the data on child sexual abuse. According to the CDC, CSA is a significant but preventable public health problem.^3^ CSA refers to the involvement of a child (person less than 18 years old) in sexual activity that violates the law and that he/she:

- does not fully comprehend,
- does not consent to or is unable to give informed consent to, or
- is not developmentally prepared for and cannot give consent to.

Although estimates vary across studies, the data shows:

- About 1 in 4 girls and 1 in 13 boys experience CSA at some point in childhood.
- 91% of CSA is perpetrated by someone the child or child’s family knows.
- The total lifetime economic burden of CSA in the United States in 2015 was estimated to be at least $9.3 billion, although this is likely an underestimate of the true impact of the problem since CSA is underreported.

Experiencing CSA can affect how a person thinks, acts, and feels over a lifetime, resulting in short- and long-term physical and mental/emotional health consequences. Examples of physical health consequences include:

- Sexually Transmitted Infections (“STIs”),
- physical injuries, and
- chronic conditions later in life, such as heart disease, obesity, and cancer.

Examples of mental health consequences include:

- depression and
- post-traumatic stress disorder symptoms.

Examples of behavioral consequences include:

- substance use/misuse including opioid misuse,
- risky sexual behaviors, meaning behaviors that could result in pregnancy or STIs such as not using condoms or other contraceptives, or sex with multiple partners, and
- increased risk for suicide or suicide attempts.

Another outcome commonly associated with CSA is an increased risk of re-victimization throughout a person’s life. For example, recent studies have found:

- Females exposed to CSA are at 2-13 times increased risk of sexual victimization in adulthood.
- Individuals who experienced CSA are at twice the risk for non-sexual intimate partner violence.

Simply put, CSA is a scourge on our society. While punishing violations is useful, preventing them would be even better.

## IV. Background: CSA Prevention and the System for Reporting

As a preliminary matter, it is important to understand that in fighting crime, there are two conceptually different strategies. One strategy is focused simply on ensuring that crime is detected and punished. On its face, with no more, this is considered good in that it is just. It might have additional benefits in the sense of locking someone up to prevent them from doing other harm, and deterring others from doing harm. But first and foremost, it is about justice and is driven by our legal system.

Prevention, on the other hand, is focused on trying to stop the crime from ever happening. Putting lights on street corners and locking doors can prevent crime. Prevention is obviously better than enforcement because it is always best to stop the harm from ever happening, rather than merely punishing a perpetrator afterward. Many of the strategies around prevention do not involve the legal system at all. Prevention often involves public works or other interventions.

While our goal here is to figure out whether the CSA educational laws help with prevention, we are incidentally interested in whether they also help with enforcement. Teaching a child how to avoid being sexually abused is best. But catching the person who abused a child is the next best thing.

Increases in reported cases of CSA can lead to improved enforcement. Improved enforcement may ultimately lead to deterrence or simply removing perpetrators from society so they can do no further harm. It becomes a form of prevention.

Whichever the mechanism, our goal is to measure whether the school-based programs that follow these CSA educational laws ultimately reduce the number of CSA cases, because that is the most important metric for doing good.

### A. The child abuse reporting process

Before laying out our quantitative strategy, it is important to provide a little background on the system of reporting and substantiating cases of CSA. Throughout the country, each state has its own reporting mechanism through which children and adults can report what they believe to be CSA. Initially the information shared with the government through a hotline or otherwise is called a “referral.” Those referrals are screened to assess generally whether they concern child abuse or neglect, have enough information to be actionable and are appropriate for the department of child protective services, as opposed to another agency. If the initial reviewer determines that those basic conditions are met, the referrals are “screened in” such that they are then deemed a report. The significance of this is that so-called reported cases of CSA have actually already undergone an initial screening to make sure they are appropriate for further investigation. Nationwide, about 45% of all referrals get screened out at this stage.^4^

Those reports are then handled through an administrative process typically involving caseworkers in the state’s child protective services organization that seek to collect evidence for the purpose of evaluating whether or not a given report is “substantiated.”^5^ While there is significant variation state to state, the national average for a given state over the years 2005 through 2019 is approximately 6000 reports of CSA a year, of which about 1200, or 1/5, get substantiated. If a caseworker substantiates a report, the report moves to a more formal investigative process that involves law enforcement personnel. Substantiation is not the same as a guilty determination, but closer to a probable cause determination.

### B. Challenges in the questions analyzed

#### 1. True variable of interest is not quantified

What we truly care about is the rate of actual CSA crimes committed in the United States. But we will never know that number, either historically or for that matter in the future. Instead, what we do know is the rate of reported CSA.

The prevailing wisdom is that cases of CSA get both underreported and over reported. They are underreported in that it is generally believed that many more children suffer CSA than actually file reports. Estimates vary, but some researchers suggest that the *vast majority* of CSA cases never get reported.^6^ CSA is underreported for many reasons, including:

- Children may not know that what is being done to them is wrong
- Children may be too traumatized, or feel shame, that keeps them from reporting what to them is an uncomfortable experience
- Family and other pressure may be brought to bear on the child not to report, either out of ignorance, concern for social stigma, or worse.
- The perpetrator might directly threaten his victim not to report it.^7^

The over reporting is more obvious in the sense that, as already observed, the national average is about 6000 reports per state per year of which only about 1200 get substantiated. “Over reporting” may or may not be the right term here, because there are at least two distinctly different reasons for that differential.

1. There may have actually been a case of CSA, but the state officials were not able to substantiate it for lack of evidence. As should be obvious, the legal system is far from perfect. Given our core value that someone is innocent until proven guilty, it is not infrequent that the guilty are not punished. We deliberately err on the side of not punishing the guilty in the absence of compelling evidence so as to reduce the risk of punishing the innocent. Or,
2. The accused may have truly been innocent. A child or an adult may inappropriately characterize what happened as CSA. Maybe it was another form of child abuse. Or maybe it was nothing. Whatever it was, in this instance it was not CSA.

This theme of actual rate of CSA versus reported rate of CSA plays out throughout this analysis.

#### 2. The two desired effects of the law we wish to measure cancel each other out

A law that requires or encourages schools to teach children and sometimes adults how to appropriately identify and report CSA in theory has at least two different beneficial effects that move in opposite directions. The first beneficial effect is to increase the reporting of CSA, because children and adults know what it is and how to report it.

But other potential benefits of the law might reduce the number of CSA crimes committed. For example, these educational programs might teach students self-protection skills such that they could better remove themselves from a potentially harmful situation. Further, these laws might act as a deterrent to discourage predators from committing CSA.

While some CSA is committed by pedophiles with psychological disorders, much of CSA is committed by mere opportunists, people who can potentially be deterred. According to an analysis of CSA offenders by the US Department of Justice, “regressed child sexual abusers prefer social and sexual interaction with adults; their sexual involvement with children is situational and occurs as a result of life stresses.”^8^ At a more statistical level, “most [people who commit CSA] are not pedophiles. In fact, about half of all victims are post-pubescent, ranging in age from twelve to seventeen, so that most of their offenders would not qualify as pedophiles. Moreover about a third of offenders against juveniles are themselves juveniles (an even larger share of the offenders against young juveniles are juveniles). These young offenders are also not pedophiles, but include a mixed group of generally delinquent youth and youth who engage in somewhat impulsive, developmentally transitory behavior. Even among adults who victimize children under thirteen, at least a third or more do not qualify as pedophiles.^9^

While it is hard to deter people who are true pedophiles with psychological disorders, we theorize that some opportunists – perhaps themselves juveniles – can be discouraged if the likelihood of being reported is perceived to increase. Further, perhaps their attempts can be foiled through strategies that can be taught to children.^10^

From the standpoint of measuring impact, though, we have to understand that those two beneficial impacts work in opposite directions. An increase in the effectiveness of getting children and adults to report CSA would be measured by an <u>increase</u> in reported cases. An increase in the deterrent effect plus potentially an improvement in the self-protection skills of children would manifest themselves in a <u>decrease</u> in the number of CSA cases to be reported and substantiated, at least over the longer haul.

## V. Knowledge Upon Which We Are Building

Research in this area has told us a couple of things. First, school programs appear to succeed at teaching children and staff about how to identify and report CSA. Indeed, this realization led many schools, well before the subject laws were enacted, to teach this topic. Second, child abuse in general, and CSA specifically, appear to be influenced by many factors from economic environments to social trends. This is important, because in our analysis we need to make sure that we control for other reasonable explanations for changes in the rate of CSA. In the next two sections we further explore this foundational research on the impact of educational programs and other factors affecting the rate of CSA.

### A. The Impact of Educational Programs

Educational programs directed at CSA typically include content which helps people – students and staff – recognize CSA and know how to report it. The strategy behind using educational programs is straightforward:

Accessing children directly through schools allows educators to reach children from a wide range of socio-economic and ethnic backgrounds. Targeting children at school rather than relying on parents to provide the information helps ensure children who are being abused in their homes still have access to CSA education.^11^

As a consequence, school-based programs have been widely used.^12^ Further, “[l]iterature reviews and meta-analysis indicate child-targeted programs can increase children’s knowledge, self-protective behaviors, and reporting behaviors.”^13^

For example, regarding child abuse generally and not just CSA, a 2021 study examined the overall effect of these school-based programs on 1) children’s child abuse-related knowledge and 2) self-protection skills by conducting two meta-analyses.^14^ The study found a significant overall effect of school-based programs on both knowledge and self-protection skills. The results of the first meta-analysis on children’s child abuse knowledge suggest that program effects were larger in programs addressing social–emotional skills of children and self-blame, than when puppets and games or quizzes were used. The second meta-analysis on children’s self-protections skills revealed that no particular individual components or techniques were associated with increased effectiveness. While the studies covered the range of child abuse, many of them focused on CSA.^15^

That seems to be as far as the research has gone. There does not seem to be much research either on:

- The extent to which schools are currently using this type of approach, except for noting that the programs now seem to be “widely-used.”^16^
- The quantitative impact of the school programs on the rate of CSA, whether resulting from the state laws or not.^17^

Assessments of the quantitative impact of school programs have been limited in their conclusions because they have largely focused on extrapolating from small sample surveys. For example, in a 2000 study, 825 women undergraduates from a New England state university filled out a survey on “sexual experiences” for research credit.^18^ Respondents were asked detailed questions regarding past histories of CSA and participation in school-based prevention programs during childhood. Sixty-two percent of the sample reported having participated in a “good touch-bad touch” sexual abuse prevention program in school. Eight percent of respondents who reported ever having had a prevention program also reported having been subsequently sexually abused, compared to fourteen percent of respondents who did not ever have a prevention program.

We hope to pick up where such research left off, and quantitatively assess the impact educational programs that follow legislation on the rate of CSA.

### B. Other Factors Influencing the Rate of CSA

There is limited research on the societal factors that may lead to CSA. Understanding those factors allows us to identify potentially confounding variables that we should control for in our analysis. Fortunately, we do not need to control for all such factors because, as explained more below, we are using a difference-in-differences approach comparing the experiences of states with different laws. We therefore only need to concern ourselves with factors that we can expect to be different between states.

In our research, we found that the Institut national de santé publique du Québec, or in English the Center of Expertise and Reference in Public Health of the Quebec National Institute of Public Health, produced a useful summary of the research on this topic. The following grouping of factors is largely from that summary.^19^

- Individual factors

Certain individual factors have been associated with an increased likelihood of being sexually abused as a child. The most consistently reported factors include: being female, being between the ages of 6 and 11 (for intrafamilial sexual abuse alone), being between the ages of 12 and 17 (for extrafamilial sexual abuse alone), having experienced physical or sexual abuse in the past, and having special needs (e.g., handicap, intellectual disability, chronic illness, and mental health problems).

- Relationship/family factors

Certain relationship factors have been associated with an increased risk of being sexually abused as a child. The most consistently reported factors include: limited supervision by parents, use of drugs and alcohol by parents, having parents with mental health problems, and being in a family where the mother’s spouse is not the child’s biological father (i.e. a stepfather family).

- Community factors

Community factors associated with an increased likelihood of being sexually abused as a child have been studied to only a limited extent thus far. However, some studies suggest that tolerance of sexual abuse and weak sanctions against sexual abuse within a community play a role in raising the risk.

- Societal factors

Various societal factors have been associated with an increased risk of being sexually abused as a child, in particular: hyper-sexualization of young people in a society, a history of denial in a society that CSA occurs, traditional norms regarding gender roles, the presence of an ideology of male sexual entitlement, weak legal sanctions against CSA, and social norms that support sexual abuse.

For the current analysis, mostly we will focus on economic data that might indicate stressful home environment differences across states, in line with the relationship factors discussed above. These would include unemployment and income levels.

### C. The Rate of CSA Is One Factor in the Adoption of the Laws

Not surprisingly, research suggests that the adoption of these educational laws is in some way causally connected with the rate of CSA cases in the particular state.^20^ Indeed, that is true of most laws. Most laws are a response to the severity of the problem they address. We explain that here because it means that the law itself cannot be an instrumental variable, which affects our choice of methodology. An appropriate instrumental variable cannot be causally connected to the ultimate outcome we wish to measure.

## VI. Hypotheses

### A. How we address the opposing forces

Since they oppose each other as discussed above, if we do not know the strength of either factor, we cannot separate out the impact of either factor. Indeed, in theory, there could be tremendous benefit in the legislation if: 1) the ability of the children increased dramatically to identify and report CSA, while 2) potential opportunists are significantly deterred from committing CSA. However, the net effect of those improvements could be, in theory, *a level of CSA reporting that is similar to before the law was enacted.* However, it seems to us more likely that one of these factors might predominate over the other, and thereby lead an overall change in the reported and substantiated CSA cases, either higher or lower depending on which effect is larger.

A final scenario we contemplated was that the number of reported cases might go down while the number of substantiated cases might go up. We could anticipate seeing that outcome if the deterrent and self-preservation benefits of the law reduce the actual number of CSA cases by a small amount, but the increase in reporting is more substantial *and* the education about proper reporting causes a decline in the number of reported cases that cannot be substantiated. In other words, if people know what should and should not be reported, we could imagine the ratio of reported to substantiated going down, while the volume of reporting otherwise would have gone up, together with the substantiated cases. Remember we calculated that about five cases get reported for every one that is substantiated. If the biggest change was that that number fell to say for every two cases that get reported, one is substantiated, that effect might overtake both the deterrent effect and the drive to increase reporting. This would cause an increase in the absolute number of the substantiated CSA cases while at the same time producing a drop in the number of reported cases. In small numbers, you could move from 10 reports and 2 substantiated cases to 6 reports and 3 substantiated cases. The reports come down and the substantiated goes up. Basically, the system becomes more efficient and effective by teaching people what to report and what not to report, resulting in more substantiated reports being made.

Conceptually, we hypothesized the following three scenarios where in each scenario, in different magnitudes, 1) the law is effective at causing the decline in actual CSA crimes (whether through deterrence or self-protection) and 2) the law is effective at educating people about what to report. If both turn out to be effective, the direction of the changes will be determined by the relative impact of those two effects.

What we did not hypothesize, quite honestly, is a fourth scenario where the number of substantiated CSA cases declines and the number of reported CSA cases grows, because that would necessarily mean that the education caused proportionately more unsubstantiated claims to be submitted.

### B. Framing the questions analyzed

Given those challenges, what can we measure that would be useful to policymakers in this field? While we lay out seven specific hypotheses below in section VI, in more conceptual terms, we sought to measure the following three types of impact.

#### 1. Measuring net impact

The most straightforward approach is to simply acknowledge that we cannot measure each impact of the law (i.e. deterrence and encouraged reporting) and simply look at the net impact the CSA educational laws have on the reported and substantiate CSA cases. While this will not tell us what we most want to know (i.e. how much the law deterred actual crime and how much the law caused previously unreported cases to be reported), the net impact on both reported and substantiated cases is useful information.

#### 2. Measuring the change in the quality of the reports

We theorized that we can measure the impact of the laws on the quality of the reports filed by examining the ratio of reported cases of CSA over substantiated cases of CSA. A detrimental direction would be an increase in that ratio, suggesting that more cases are being reported that ultimately are not substantiated.

### C. Specific hypotheses

We have a total of seven hypotheses and explain each in turn. In developing the five, in addition to CSA, we examine, in a similar manner, whether the enactment of the law is predictive of the rate of child abuse generally, not just specifically CSA. The theory is that the educational programs teach children to be more vigilant generally, and those skills can be used in other areas of child abuse as well.

Here are our alternative hypotheses based on those theories:

1. Educational programs on CSA that follow state enactment of laws requiring or encouraging such programs impacts the number of reported CSA cases.
2. Educational programs on CSA that follow state enactment of laws requiring or encouraging such programs impacts the number of substantiated CSA cases.
3. Educational programs on CSA that follow state enactment of laws requiring or encouraging such programs impacts the number of substantiated <u>Child Abuse (“CA”) cases</u>, as opposed to simply CSA cases.

The ratio of reported cases of CSA to substantiated CSA cases is an important justice metric, because it reveals the accuracy of the reports. Reports of CSA that cannot be substantiated cause harm to the one accused. Therefore, one potential benefit of the legislation is improving the accuracy of the reports, meaning that a greater share of the reports lead to cases that can be substantiated.

We also need to make sure that any change in this ratio metric is not due to structural reasons that limit capacity to substantiate reports, such as a fixed number of social workers that has reach capacity. We can test this by studying the number of incomplete investigations.

The final two hypotheses are:

4. Educational programs on CSA that follow state enactment of laws requiring or encouraging such programs decrease the ratio of reported CSA cases to substantiated CSA cases.

5. Educational programs on CSA that follow state enactment of laws requiring or encouraging such programs do not affect the number of incomplete investigations of reported CSA cases.

Our theory is that teaching kids and some adults how to accurately identify CSA and appropriately report it should produce more accurate reports. Historically, only about 1/5 of the CSA cases reported lead to a determination of “substantiated” report (which of course is due to many factors, and many of the reports are appropriate even if they case worker is ultimately not able to substantiate the report). As a consequence, there is ample room to improve on the quality of that reporting.

The null hypotheses are those same hypotheses but concluding that there was no significant difference, or at least not the significant difference we hypothesized.

### D. Outcomes we theorized away

We feel comfortable assuming away certain potential negative outcomes, such as:

- The CSA educational laws reduce the deterrent effect. To do this, the laws would need to somehow make it less likely that cases of CSA would be punished. We do not think this likely, so we will assume this does not occur on average over the 50 states.
- The CSA educational laws cause real cases of CSA to go unreported when otherwise they would have been reported. If the CSA educational programs in schools are poorly designed, in theory they might cause confusion or might actually lead someone to misunderstand and <u>not</u> report a case they would have otherwise reported. While it is not impossible that a particular school program could be defective enough to cause that outcome, we are making the weaker assumption that on average, these school programs across the country would not reduce the reporting of real cases.

### E. Our use of the concept of statistical significance

Our primary audience for this paper is policymakers. In many scientific papers, including those in data science, the authors adopt the threshold of a 95% confidence level before stating a conclusion. That certainly makes sense to us when a group of scientists are exploring something that they can label a truth.

Policymakers operate in a different world, where they are called to make decisions all the time, and need to make use of whatever information they have available to them. If a given conclusion is associated with only a 90% confidence level, it might still be the best available information and it might still be information on which the policymakers should rely in the absence of better information.

In this paper, we take the strategy of presenting estimated conclusions, and rather than simply limit those conclusions to only those which satisfy the 95% confidence level, we provide conclusions in which we have less confidence, but we state what those confidence levels are. We may state it either as an explicit confidence level, or the magnitude of the standard error. That way policymakers can decide for themselves whether they wish to act on the information.

## VII. Data Description

We started with a June 2021 legal analysis of the 50 states regarding educational laws directed at reducing CSA. Fortunately, we have 31 states that have adopted the treatment variable prior to June 30, 2019, which leaves 19 states for comparison purposes, as well as the other states prior to their treatment.

### Subjects

Our subjects in this study are American children, with an emphasis on those who are school-aged as the recipients of the treatment, although we will be measuring the outcome likely with regard to children of any age. In 2019, there were approximately 73 million children in the United States.^21^

### Treatment versus Control

The treatment condition will be the CSA education following a state law, and the control condition will be those who do not receive such education, or they might receive other interventions discussed below under alternative laws.

### Real World Challenges to the Research

The laws are typically in the form of requiring or suggesting that schools teach children and sometimes adults to recognize and report CSA. The challenge with this analysis is the variance in the timing with regard to when laws are actually implemented. Often a state will not appropriate money, or at least not right away, to allow the implementation to proceed in a timely way. These are referred to as unfunded mandates. Further, as already observed some of these laws do not require educational programs, but merely encourage them. That difference in the law might produce differences in the degree of implementation, i.e. how many schools actually participate, or how quickly they implement the laws.

### Strategy Attempted to Overcome that Challenge

We therefore wanted to develop a metric that reflects the degree the laws are implemented, or more specifically the number of schools implementing such programs across time. Our choice of metric was the frequency with which schools and their administrators tweet about the law. Our theory was that the more tweets we see by schools about the law, the greater the number of schools implementing the law. Unfortunately this strategy did not work out because there were an inadequate number of school tweets on the topic. So we had to proceed without that data.

### Specific Variables of Interest to Be Used In the Study

Broadly speaking, we have three groups of independent variables of interest and one response variable per hypothesis. The independent variables of interest include:

1. Variables associated with the specific type of CSA law enactment, and
2. Control variables to capture other factors that impact the rate of CSA

### Dependent/Response Variable

The response variable for each hypothesis is mined from a child abuse data set maintained by the Department of Health and Human Services (“HHS”) that tracks reported instances of child abuse. The National Child Abuse and Neglect Data System (“NCANDS”)^22^ is a data collection system that gathers information from all 50 states, the District of Columbia, and Puerto Rico about reports of child abuse and neglect. We applied to HHS for access to that database, under the oversight of the University of Michigan Institutional Review Board. The data are de-identified in the sense that we do not have the name or other personal information about the victim or the perpetrator. We have about 150 variables with regard to each incident.

For our actual Y value, we will use 7 different ones for the 7 hypotheses. Specifically, we will use the following as alternative Y values:

1. Level of reported cases of CSA
2. Rate of substantiated cases of CSA
3. Rate of substantiated cases of child abuse generally, not just CSA
4. The ratio of reported cases of CSA to substantiated cases of CSA (i.e. by state by year, total reported divided by total substantiated)
5. Number of incompletely reviewed cases by social workers

We wanted to understand if the impact that we might see on the rate of substantiated CSA cases has something to do with the resources available to state level to handle an influx in newly reported CSA cases. Our hypothesis was that if there were resource constraints, we would see an increase in the number of cases that were reported out of the end of the year as incomplete, meaning that no conclusion had been reached.

### Independent Variables

We have three categories of independent variables and in the following sections we identify the specific independent variables to be used in each of those three categories.

### CSA Educational Law Enactments

Not every CSA educational law was the same. We tracked the differences. In this category, using the existing legal analysis, we have 5 variables we plan to use in the regression. Those include:

1. The effective date of the CSA educational law
2. Whether that law was mandatory or voluntary
3. Whether that law required the education of all staff, or only some staff
4. Whether that law required the education of all students, or only some grades
5. Whether that law included an opt-out provision through which parents could remove their children from the program

### Control variables

To isolate causation of the variables of interest, we must include control variables that are known or suspected of influencing the response variable. We have two categories of control variables as follows:

#### 1. Other CSA laws

While educational laws have been adopted, over the last 10 years some states have chosen other legal strategies for addressing CSA. It is quite possible that in these other states those laws are affecting the number of CSA cases. As a result, we will control for this by including in the regression variables that track when other laws were adopted during this time period including:

a. Screening school personnel
b. Criminalizing certain behaviors by school personnel
c. Establishing taskforces to consider policy changes
d. Using posters as a more passive way of educating students

In this depiction, educational laws are the CSA educational laws that we are analyzing as our treatment. The other laws are all directed at CSA, but take different public policy approaches to trying to reduce the number of CSA cases.

**Figure 1.**
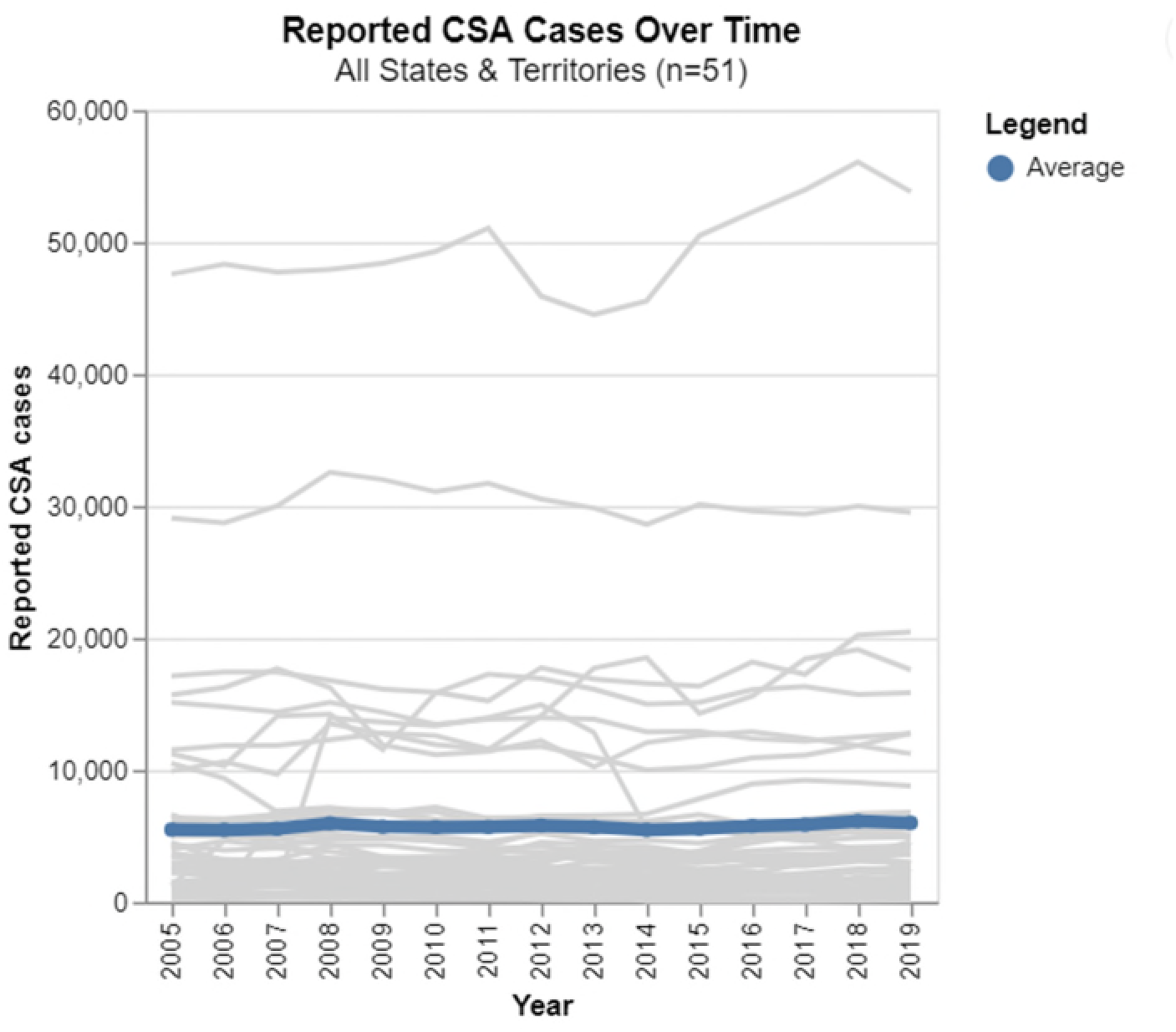

**Figure 2.**
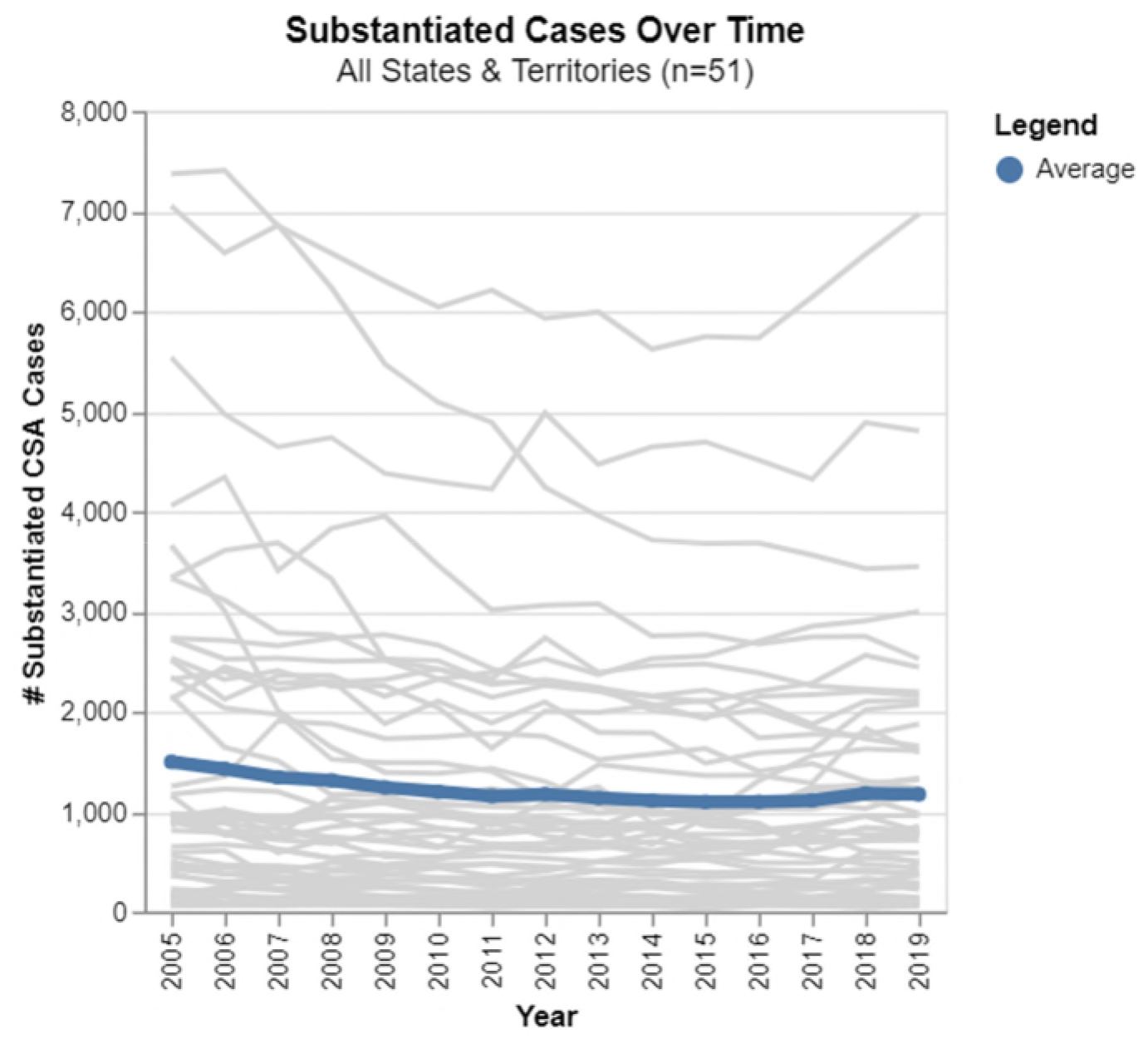

**Figure 3:**
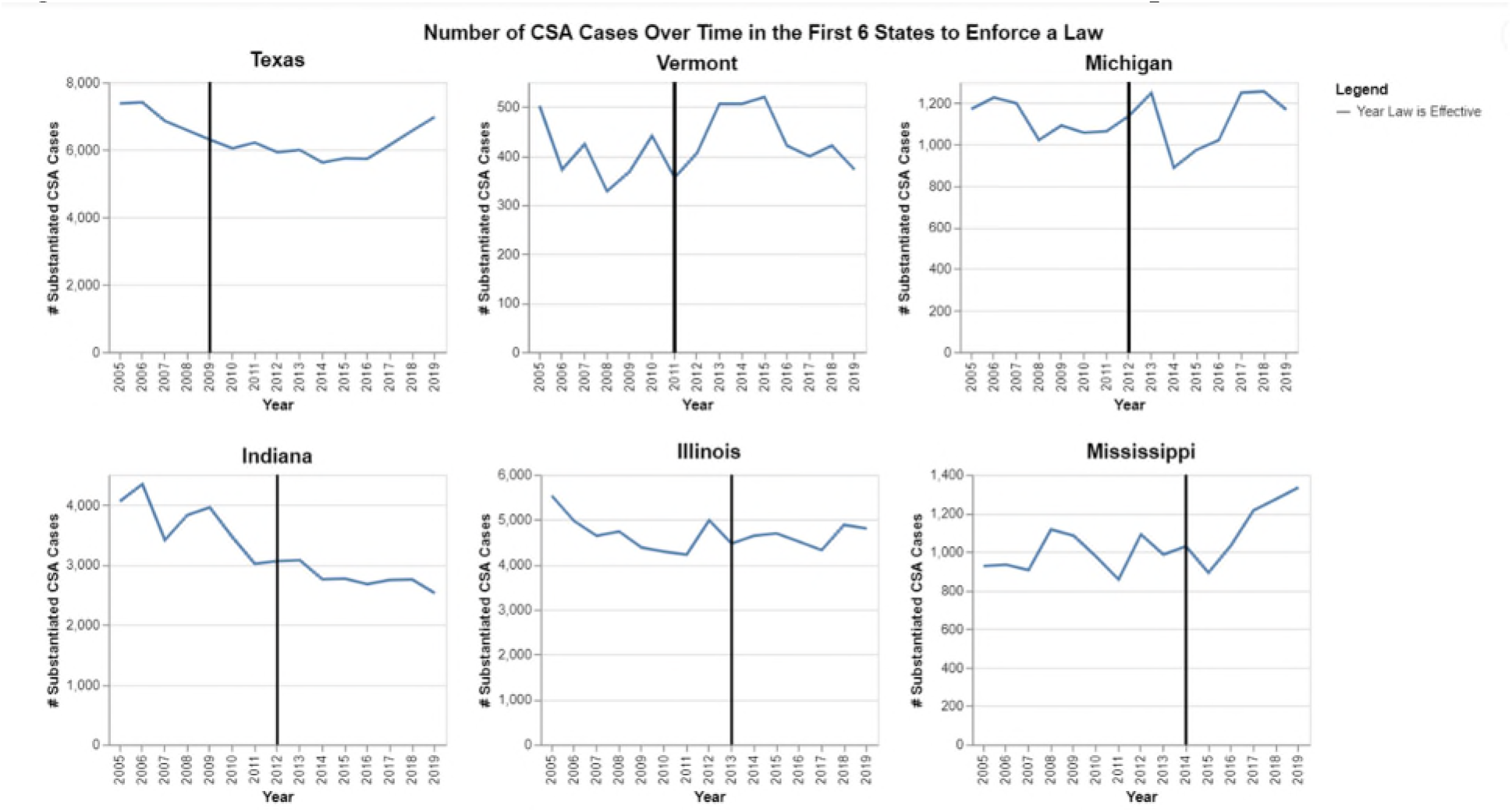
Rate of Substantiated CSA Cases for the First Six States to Adopt the Laws

**Figure 4.**
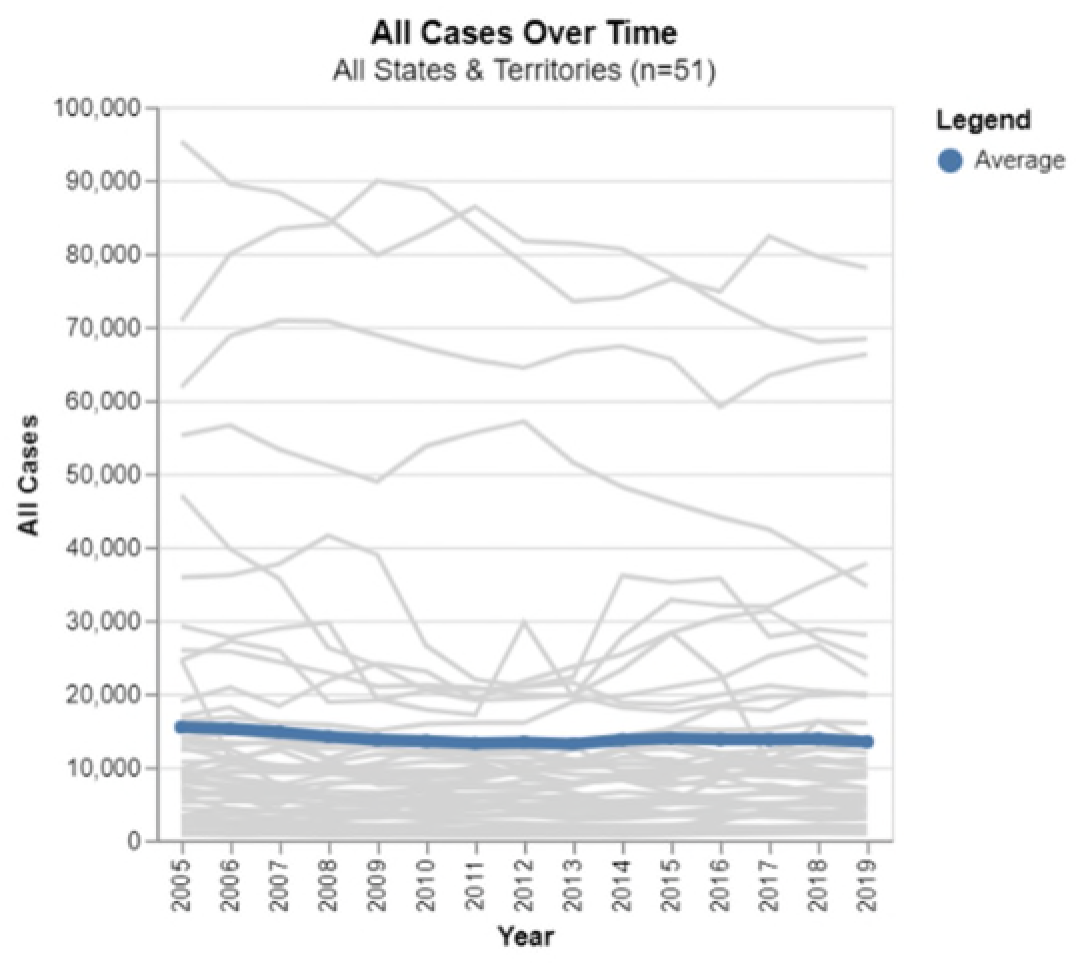

**Figure 5.**
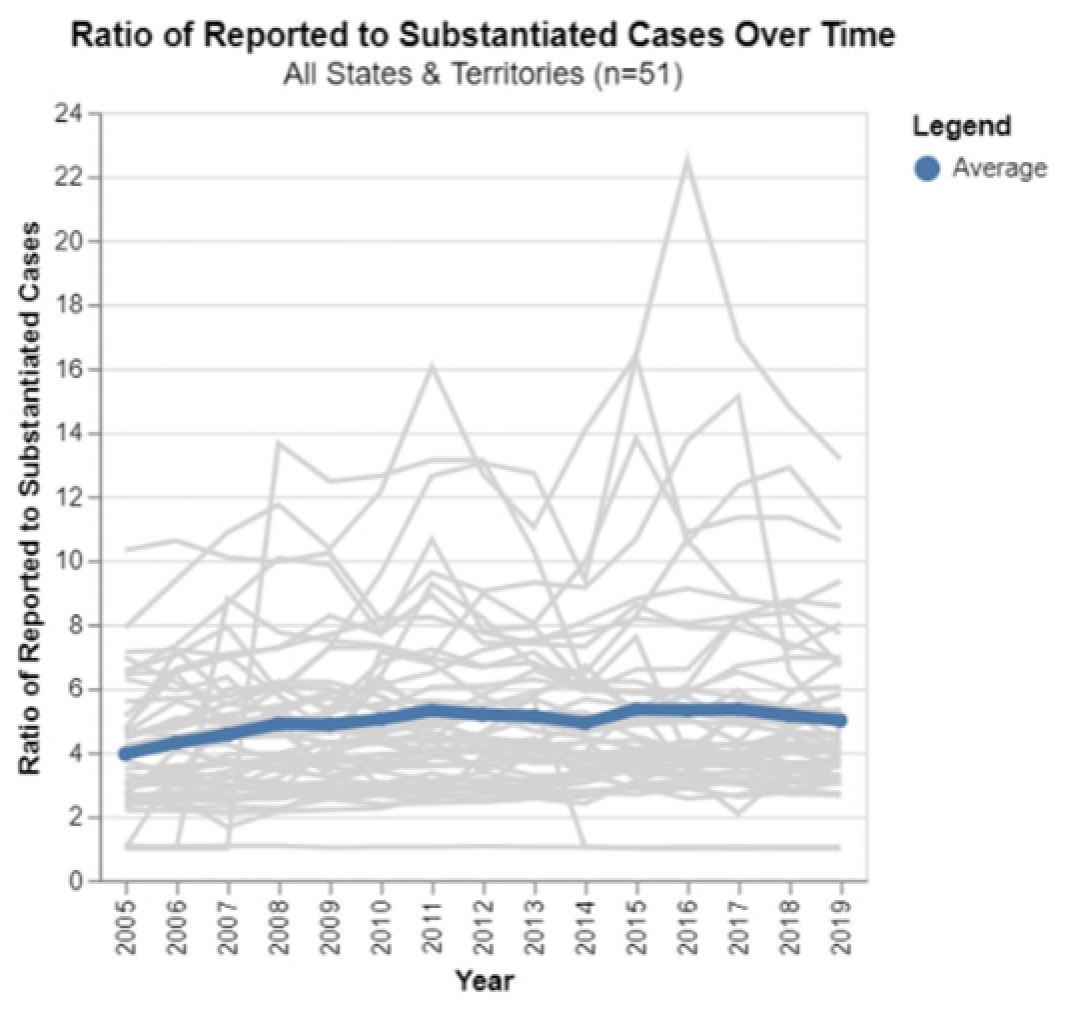

**Figure 6.**
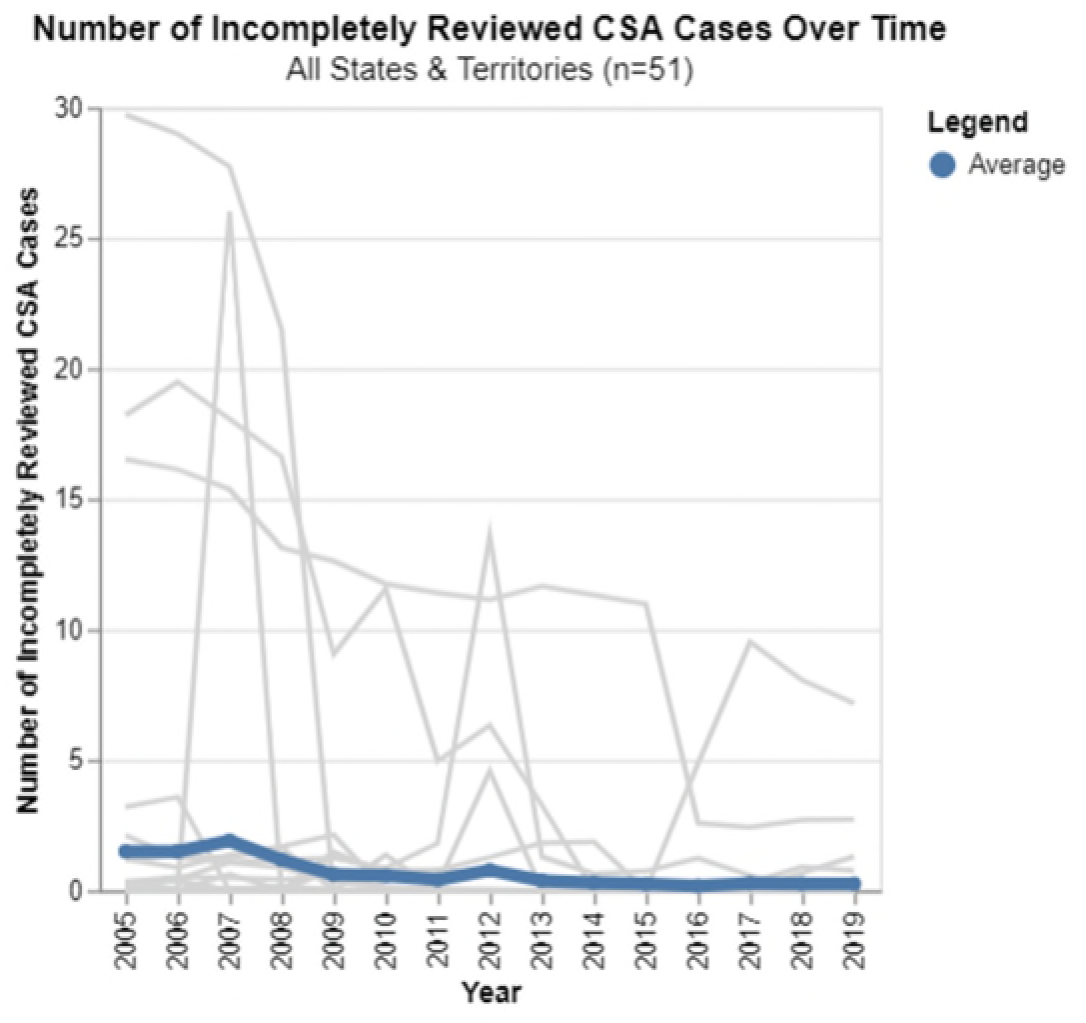

**Figure 7.**
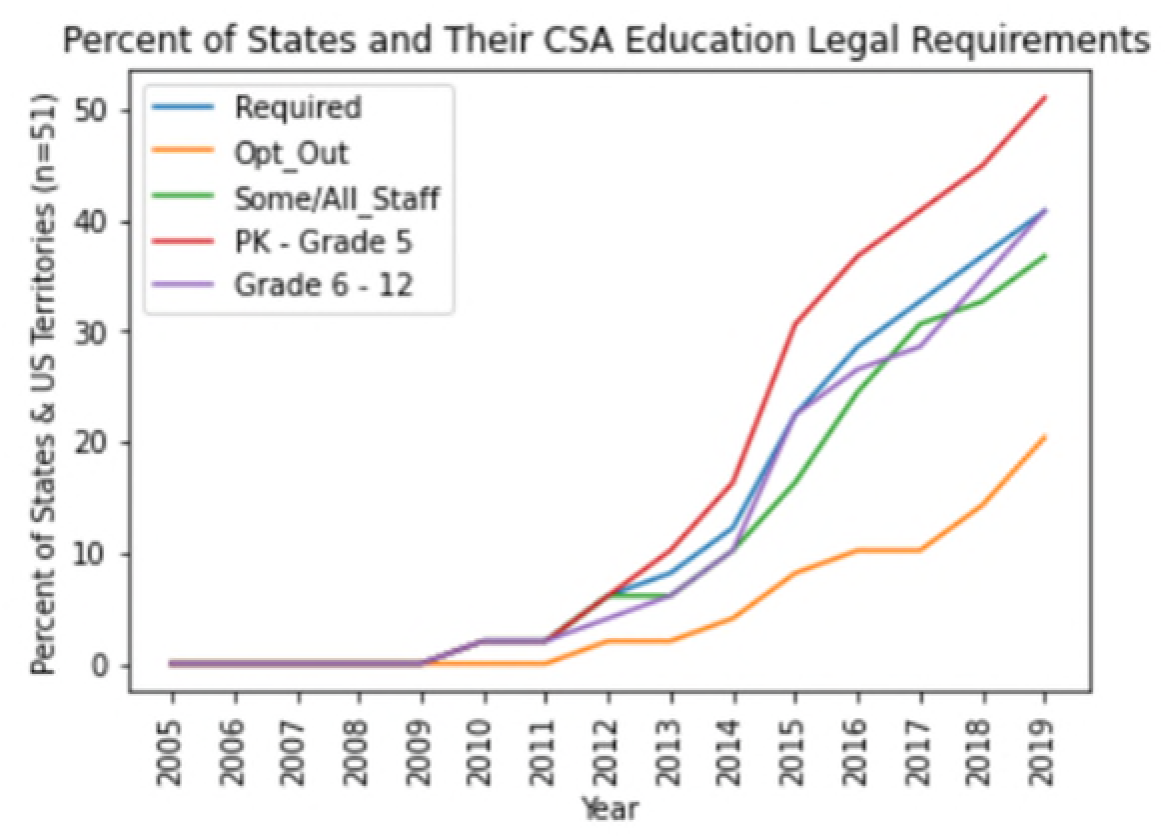

**Figure 8.**
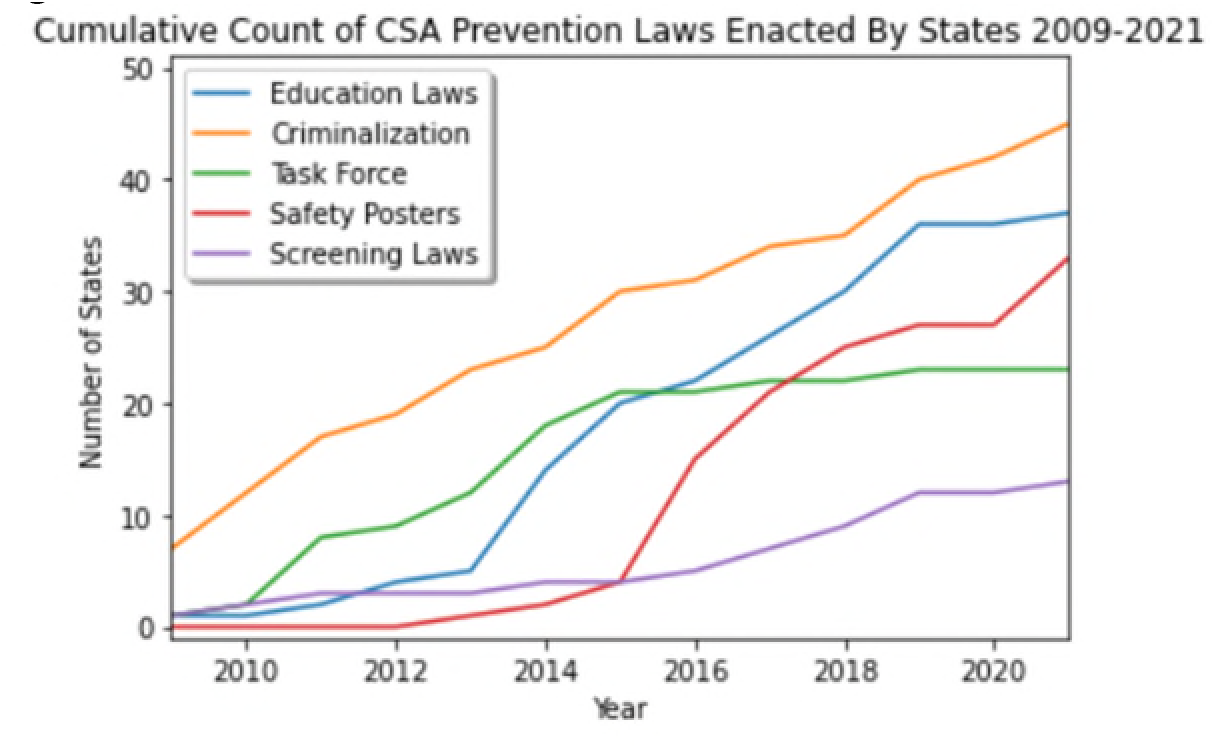

To convey the same information in a more meaningful way across the years, we have prepared an interactive visualization showing legislative enactments by year as Figure 9 at: https://bradleymerrillt.github.io/ProjectPrevent/docs/adoption_of_csa_laws.html

#### 2. Other factors known to be associated with a rate of CSA, and that vary from state to state

This will likely include the unemployment level and income levels by state. We also need population because we use that to weight the multivariate regression analysis. We obtained the data from the following sources:

- Population data: Census Bureau, State Population Totals
- Median Household Income: Census Bureau, Small Area Income and Poverty Estimates (“SAIPE”) Program
- Unemployment: Bureau of Labor Statistics

The visualizations of those three demographic data sets are included as Figures 10 (Median Household Income), 11 (Population) and 12 (Unemployment Rate) in our interactive visualizations on our colaboratory notebook here:

**Figure 10:** https://bradleymerrillt.github.io/ProjectPrevent/docs/mhi_by_year.html

**Figure 11:** https://bradleymerrillt.github.io/ProjectPrevent/docs/pop_by_year.html

**Figure 12:** https://bradleymerrillt.github.io/ProjectPrevent/docs/ur_by_year.html

**Figure 13:**
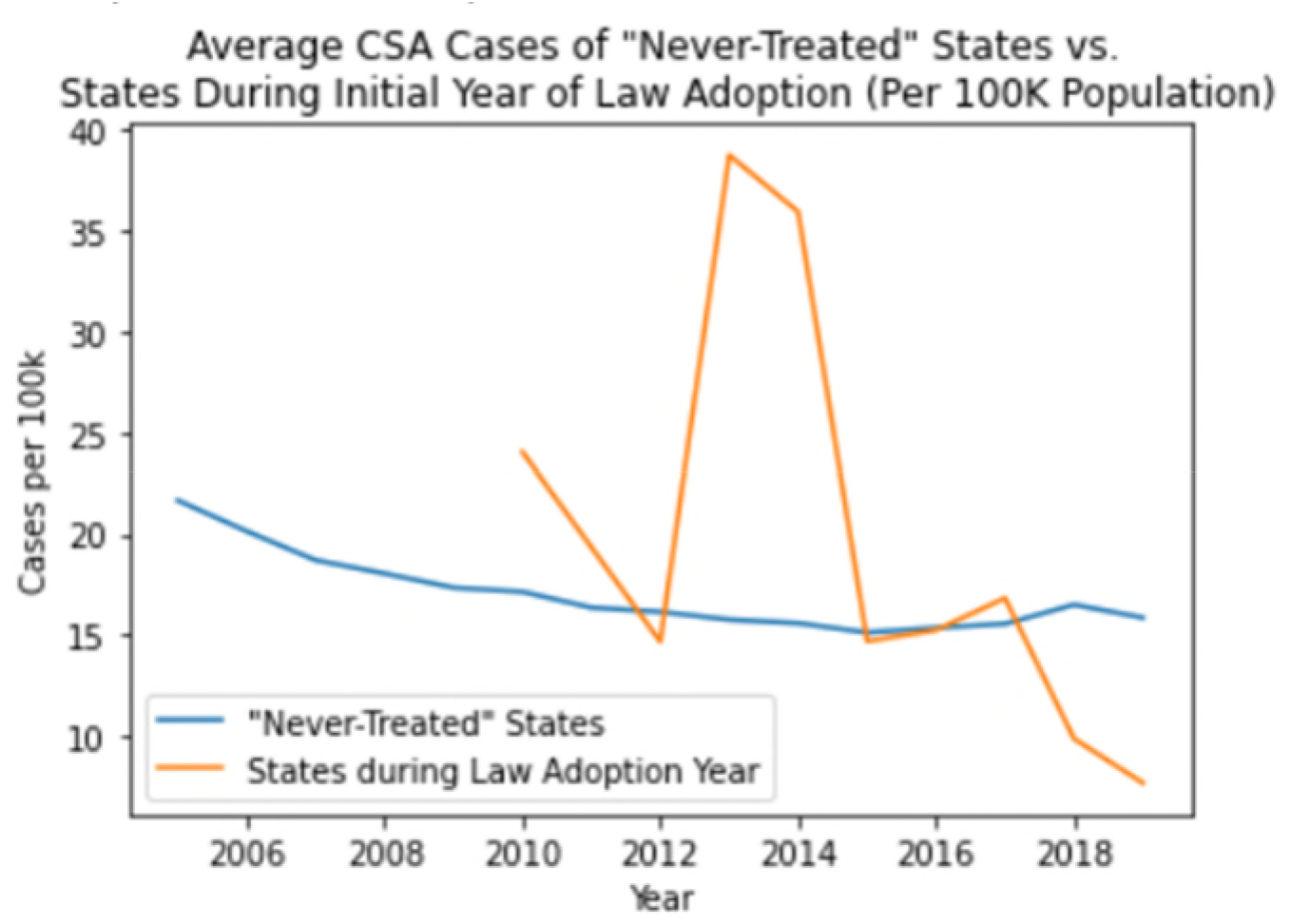

**Figure 14.**
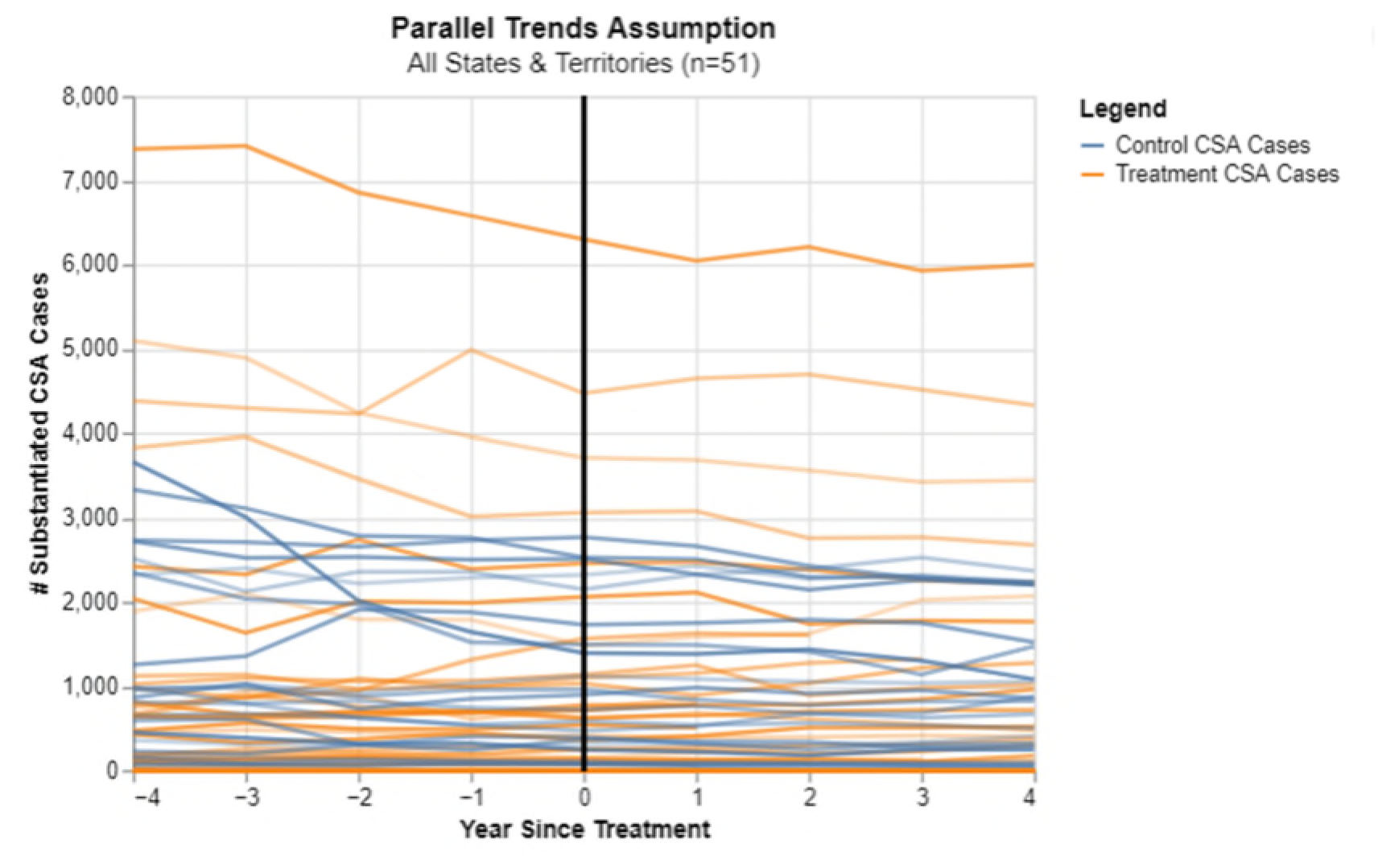

**Figure 15.**
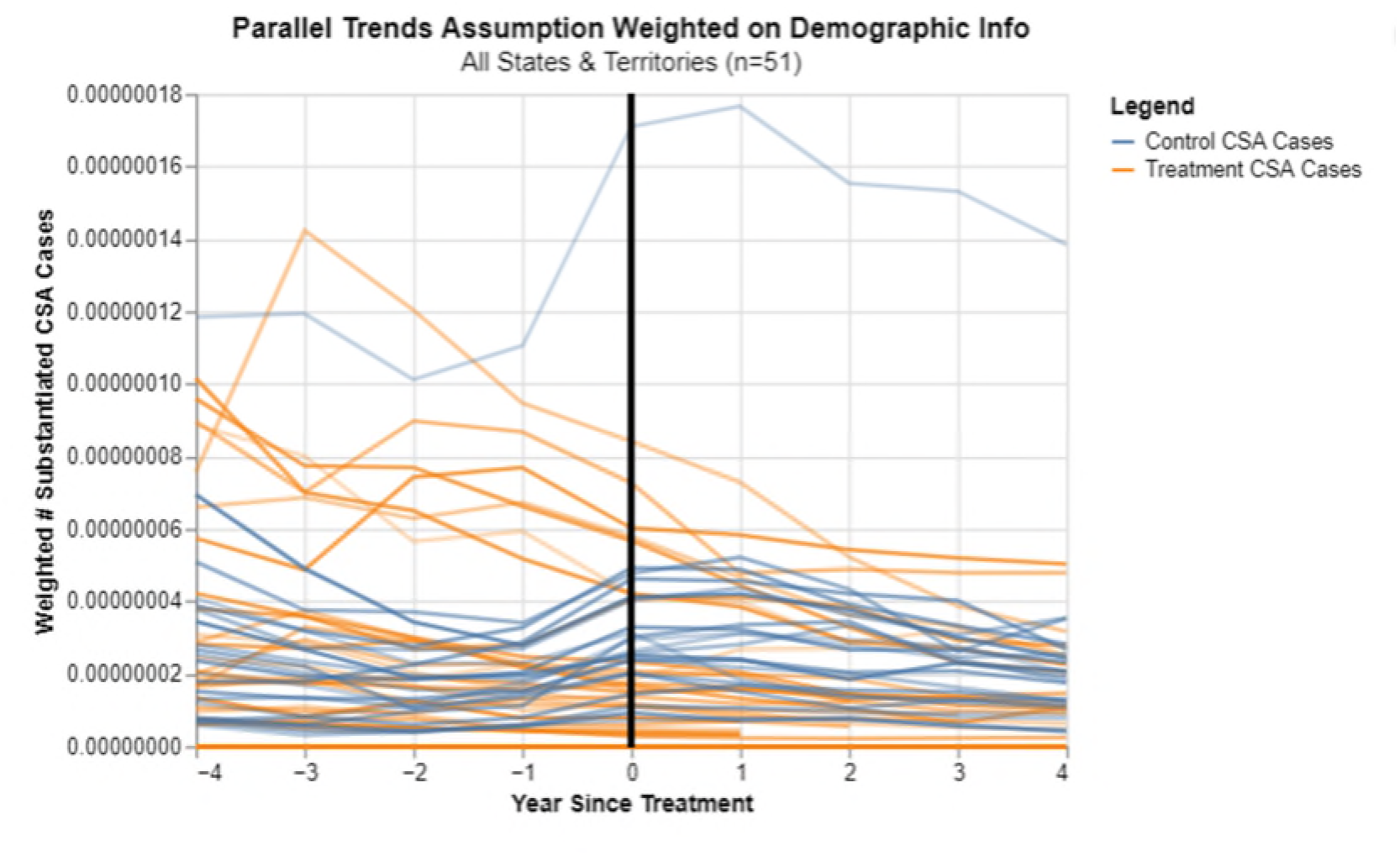

**Figure 16.**
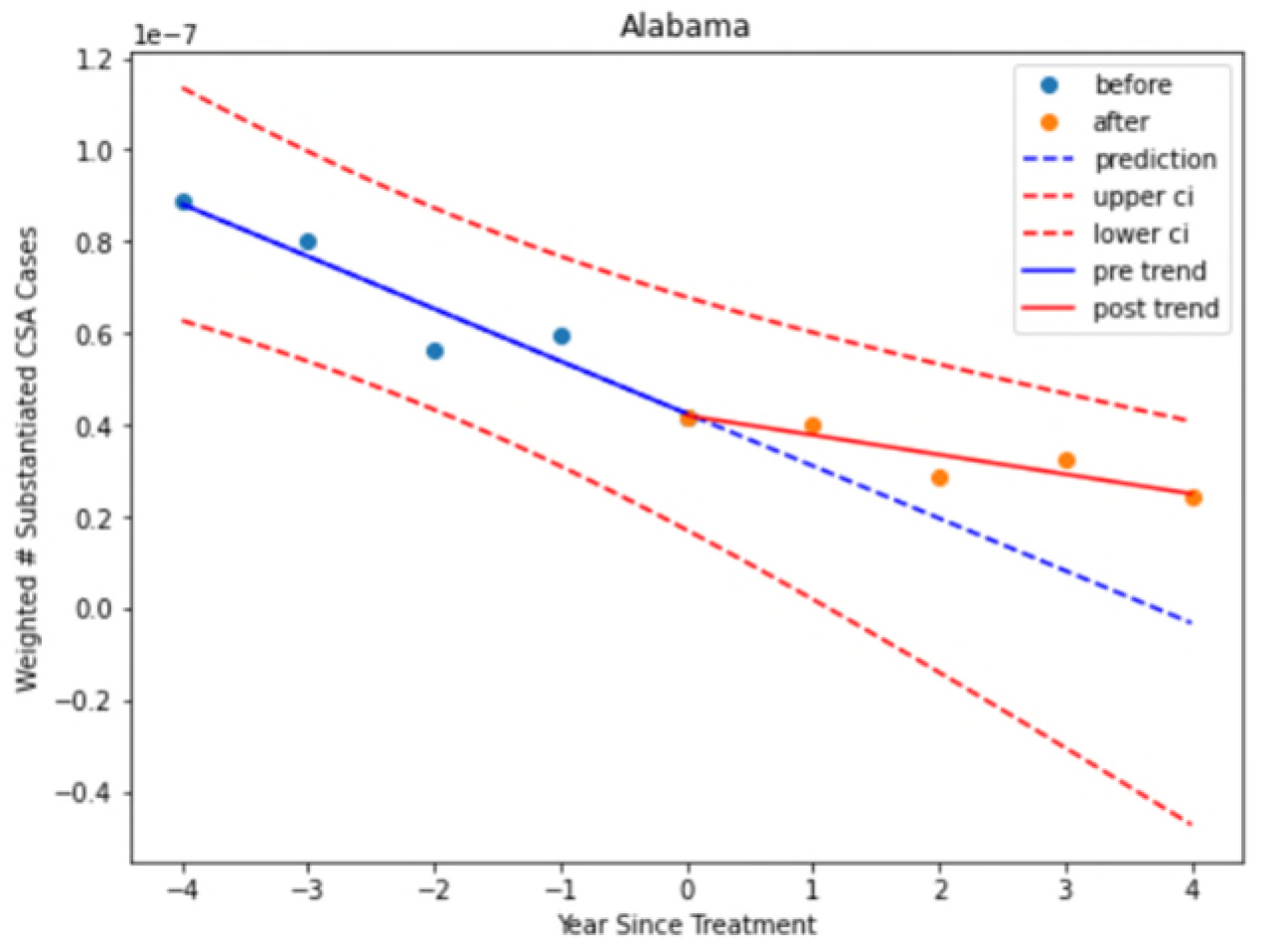

**Figure 17.**
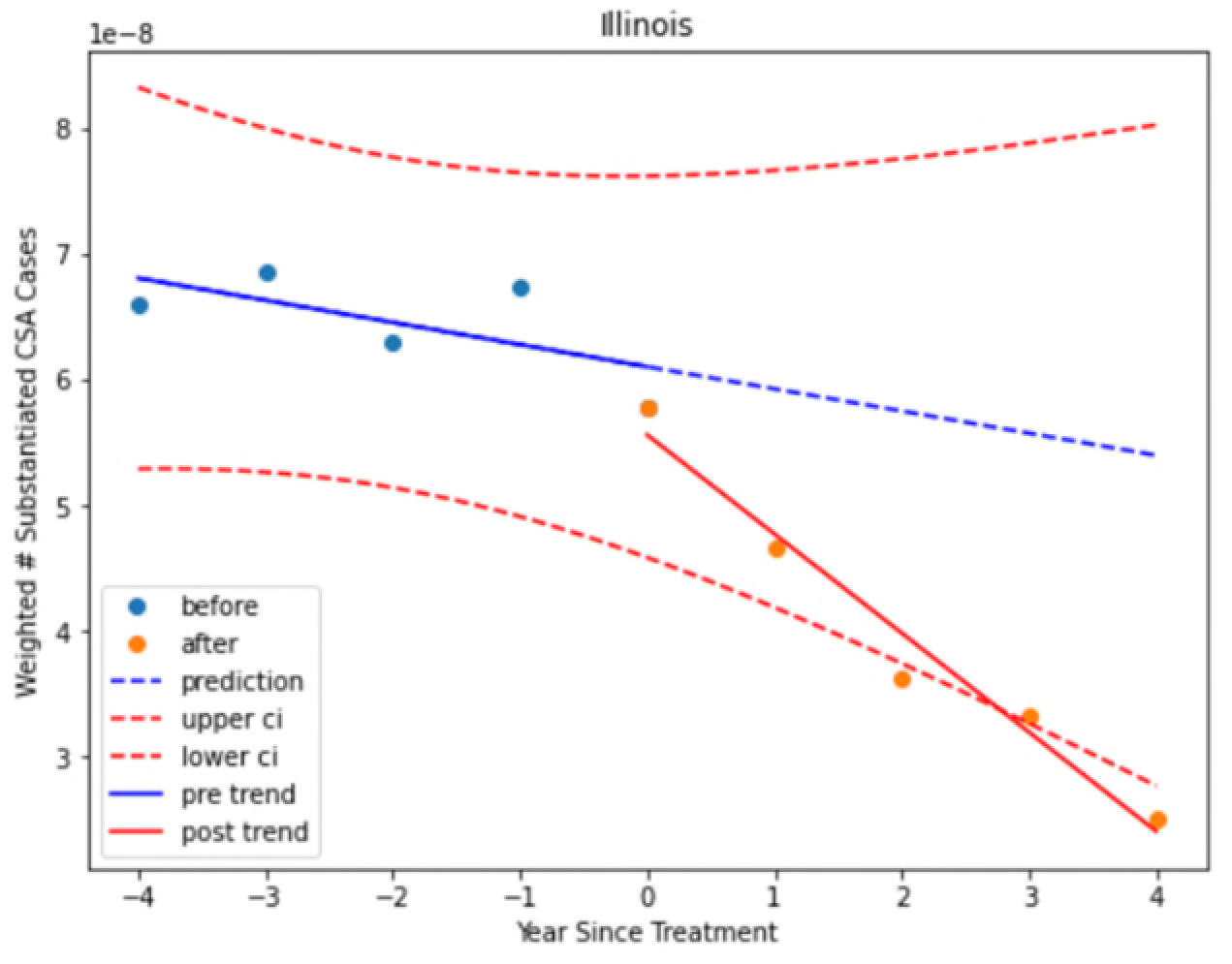

**Figure 18.**
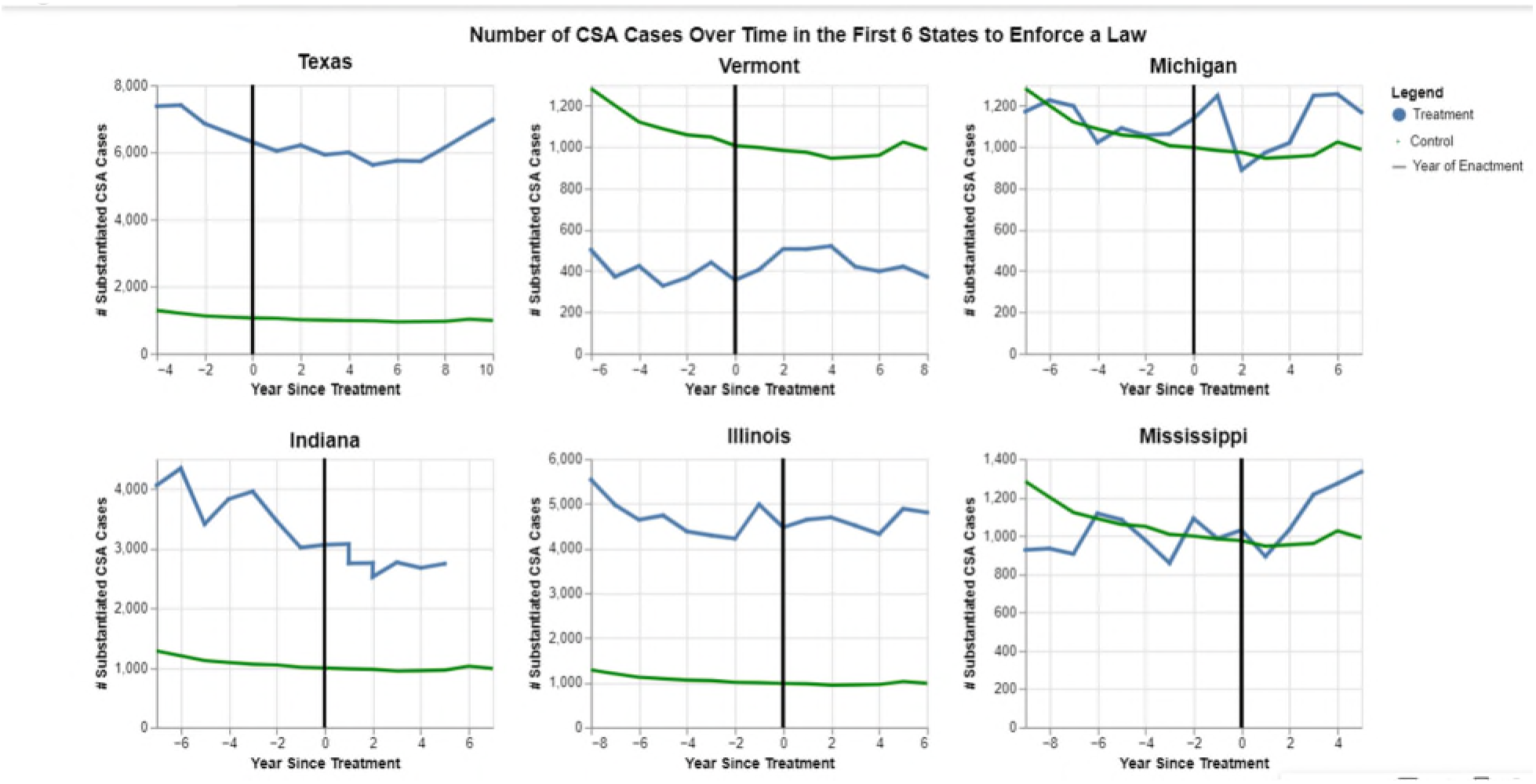

**Figure 19:**
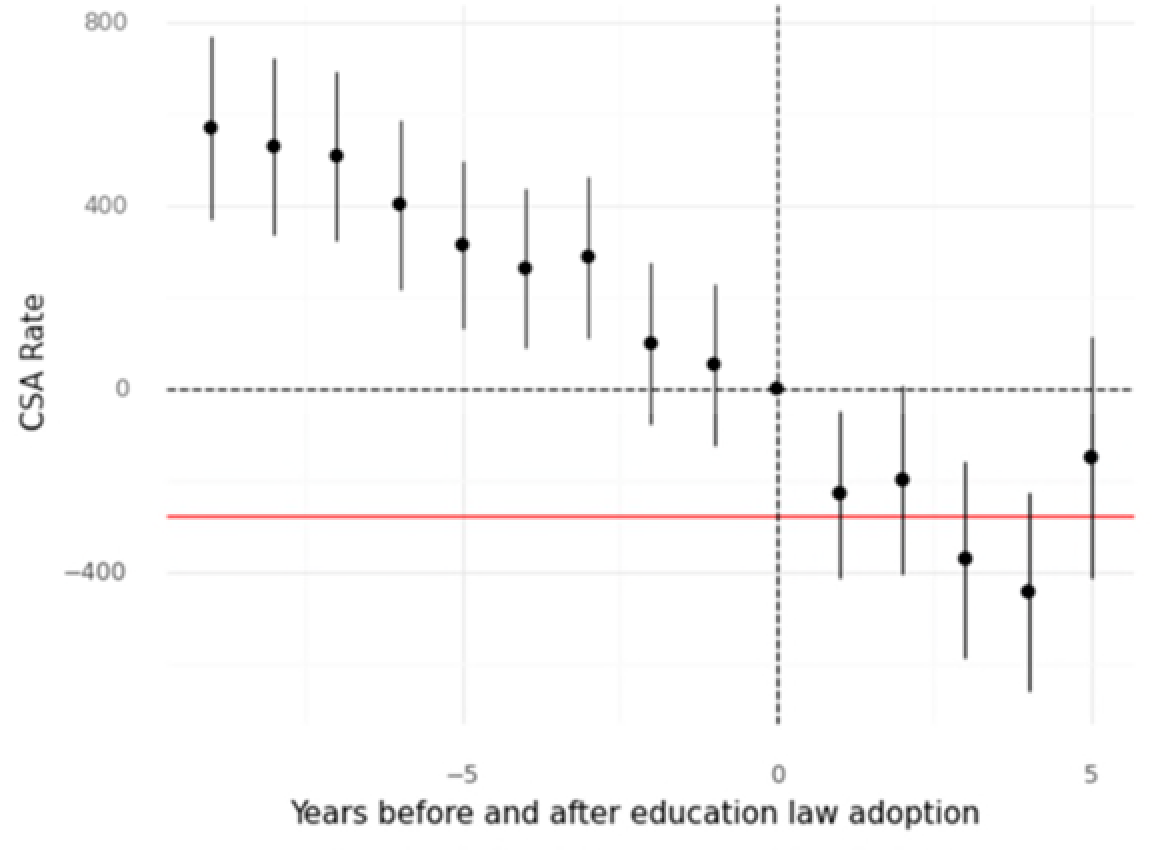
Event Study for Substantiated CSA Cases Using Regression

**Figure 20:**
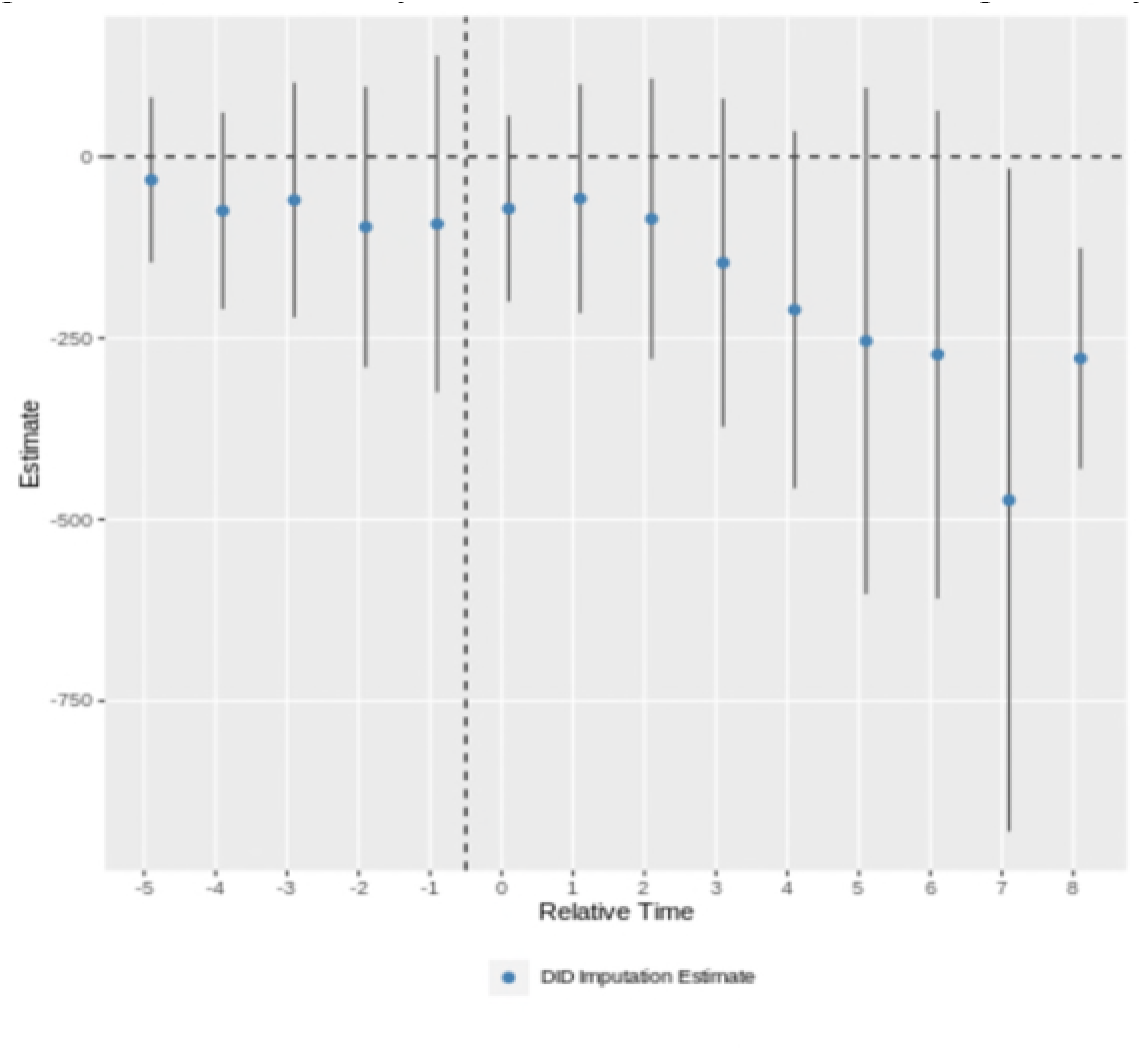
Event Study of Substantiated CSA Using Borusyak

**Figure 21.**
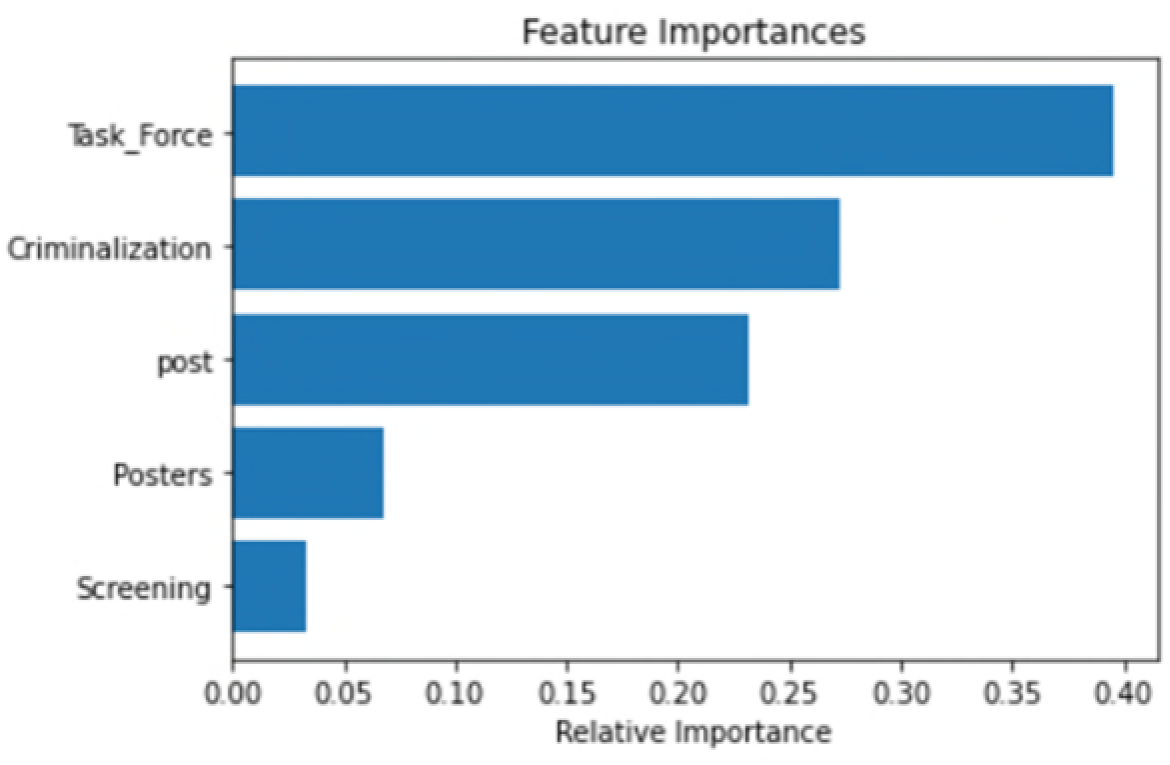

**Figure 22.**
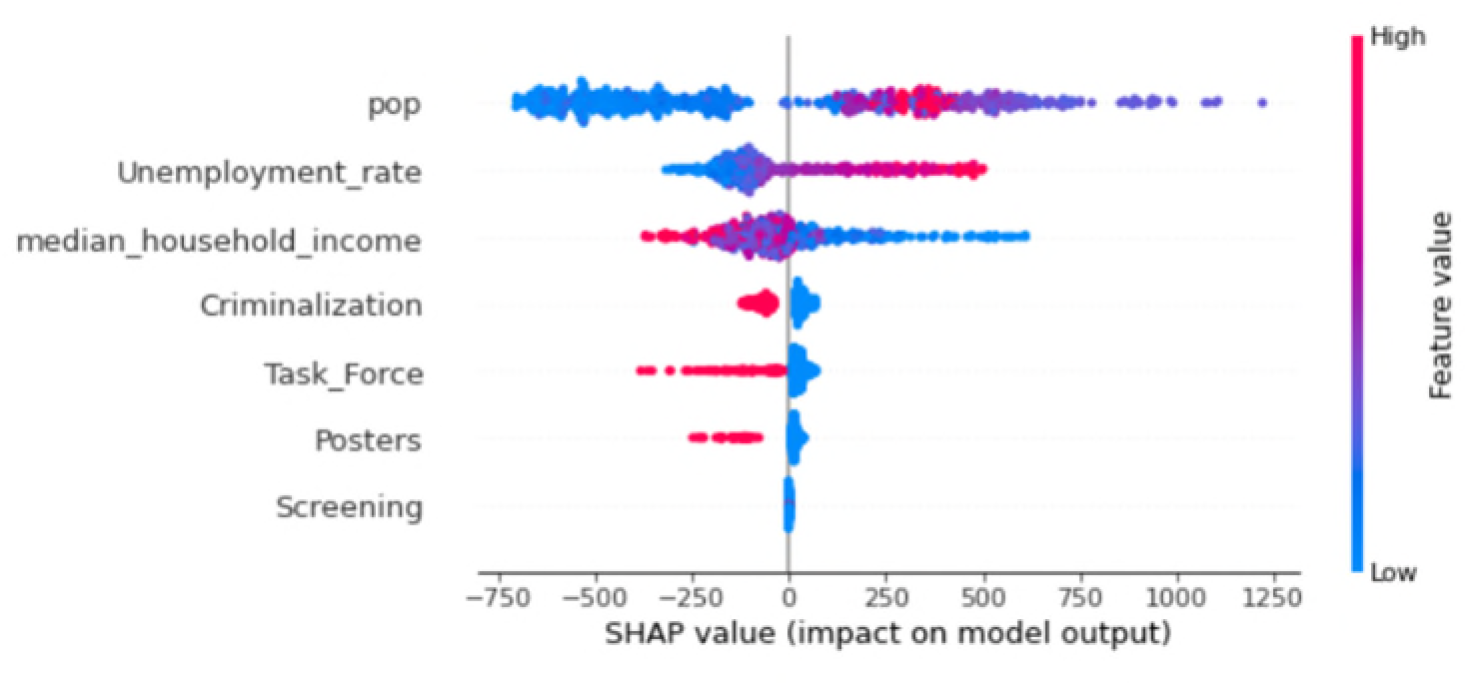

## VIII. Data Cleaning and Processing

### Legal Data Set

We use the report generated by the nonprofits entitled “A Call to Action for Policymakers and Advocates: Child Sexual Abuse Prevention Legislation in the States”^23^ as our ground truth for the legal data. We did not do independent research to confirm any of the findings. In particular, we used it not just for the rate of adoption of the CSA educational laws, but we used it to discern the key features of those laws, and to identify the adoption of other CSA legal interventions such as:

- Screening/Dismissal of School Employees to Prevent CSA,
- Criminalizing Educator Sexual Misconduct,
- State Task Forces on CSA, and
- Student Safety Posters.

We had to manually translate the data from the PDF into a machine readable form. The only data we added, which was done by the team that included an attorney, was to use the links provided in the paper to find the date on which the laws became effective. The report had only identified the date on which the laws were enacted. Since sometimes there can be a considerable delay between enactment and effectiveness, we wanted to use the effective date as we thought it was more relevant.

Apart from that research we added, we also had to make a few judgments. For Student Safety Posters, the paper discussed the fact that certain states allowed nonprofits to put up posters in schools, but for the purposes of our analysis, we did not count those instances for the legal posters column as there was no law mandating posters. However, one state, Vermont, was reported to have a regulation that required posters, so we counted that as a law. In other words, we treated both statutes and regulations as law, but not mere voluntary action by a nonprofit or others.

After finding the effectiveness dates, because we were doing an annual analysis, we decided that a year would be counted as the first year of effectiveness if the law went into effect on or before July 1 of that year. We used dummy variables to reflect the effectiveness of the laws.

### NCANDS Data Set

The data that HHS provided us was collected from the state departments of child services on an annual basis. The data set includes about 150 features for, at least in theory, every case of CSA reported each year.^24^ These data are very sensitive with regard to the privacy issues as not only do they involve children, but particularly vulnerable children. The columns we used included:

- ’subyr’- The year of data submission based on the report disposition date (The month, day, and year that a decision was made by CPS or by a court regarding the disposition of a CPS response).
- ’staterr’- the state which submitted the child file
- ’rptid’- A unique identification assigned to each report of child maltreatment. Multiple children can be included in a single report id.
- ’chid’- A unique identification assigned to each child. Child ID is unique within the state.
- ’rptfips’- The combined state and county Federal Information Processing Standards (FIPS) code of the jurisdiction to which the report of alleged child maltreatment was assigned for a CPS response.
- ’rptdt’- The month, day, and year that the responsible agency was notified of the suspected child maltreatment referral.
- ’chmal(1-4)’- The type of maltreatment that occurred. This includes physical abuse, neglect, medical neglect, psychological or emotional maltreatment, sex trafficking, sexual abuse, no alleged maltreatment, or other. These columns can also contain NA or ‘missing’ values. The four columns correspond to up to four forms of treatment that can be listed on the report.
- ’mal(1-4)lev’- The determination resulting from the CPS response to a report of alleged child maltreatment.

“Substantiated” or “Indicated or Reason to Suspect” are considered to be victims of maltreatment. For our analysis, we only considered a value of 1 (“Substantiated”) as a substantiated case.

It is an unfortunate reality that some kids are abused in multiple ways. Thus the data set allows for up to four different possible forms of abuse per case. We used any instances which included CSA, (found in any of the four ‘chmal*’ columns) and were substantiated (’mal*lev’ == 1). We then aggregated rows, grouping by state and year.

In the case of both CSA and child abuse generally, we wanted all the cases that were reported and all the cases that were substantiated. So that we could account for every case, we also wanted to keep track of those cases that were not closed for one reason or another. To get the percent of reported CSA cases which were not closed, we searched for NULL or 88.

### Demographic Data

The demographic data proved easier and had no challenges as that data is routinely used from the sources we obtained it from and required no particular cleaning.

### Data Cleaning

While the compliance is generally pretty high in NCANDS, not every state in a given year supplies the data requested to HHS. That compliance has grown over time, but during the early years there were a few states that failed to supply the data. The biggest omissions were Oregon and North Dakota in that order. Oregon failed to supply data from 2005 until 2012. Even though Oregon adopted the CSA educational law in 2015, we thought enough years were missing from the Oregon rows that we needed to drop the state entirely. North Dakota did not adopt the CSA educational law, and because it was missing the first five years, we decided to drop it as well.

On the other hand, Maryland and Michigan simply missed one year, so we filled in that year by taking the average of the two succeeding years. We did that so long as it was during a time period of mere control status, where there was no effective CSA educational law within two years.

While we had data from the legal data set on exactly which grades were affected, we thought it would be more revealing to do the analysis by putting the grades into two educational bins, basically young and older. That seem to fit the philosophy behind many of the laws. Thus, we created bins for prekindergarten through fifth grade, and another one for sixth grade through twelfth grade. We used two dummy variables to indicate whether the majority of students in the indicated grade bin were subject to the law or not.

Also on the legal data set, a few of the states such as Indiana and California amended the laws a few years after original enactment to expand their coverage, by changing the particular features of the CSA educational laws for example to expand the grades affected. We were able in our data set to then simply change the dummy variables in the year in which the amendments took place to account for the new features of the law.

To include trends across states in the weighted regression, we added data to reflect for each state for each year the series 1 through 15. So for the “Alabama trend” column, for example, the numbers went from 1 through 15 for the 15 years of Alabama rows, and zero otherwise.

## IX. Empirical Methods

As already observed, our method involved a DID analysis. The DID analysis contrasts the states that have an educational law – 31 through June 30, 2019 – with those states that do not and those prior to their own implementation.

### Method 1: Determine the Treatment Effect through a DID Regression

The regression we use is depicted as the following model:

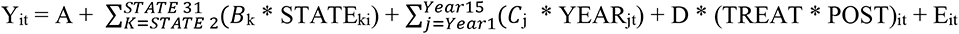

STATE_ki_ is a set of dummies for each state

B_k_ captures time invariant state fixed effects

YEAR_jt_ is a set of dummies for each year

C_j_ captures time effects that are common to all states

The regression approach tends to work well so long as there are not major heterogeneous effects among the states. Unfortunately, there were significant heterogeneous effects in the substantiated CSA case analysis, perhaps partly because there were some variations in the CSA educational laws.

### Method 2: Determine the Treatment Effect through a Difference in Differences (“DID”) Calculation that Employs Grouping Followed by Regression

The general idea of “differential timing” is discussed in Scott Cunningham’s book, “Causal Inference: The Mixtape” in the section on “Twoway Fixed Effects with Differential Timing.”^25^ One approach described in that chapter was originally explained in an unpublished manuscript in 2018 by A. Goodman-Bacon on “Difference in Differences with Variation in Treatment Timing.”^26^ In executing this approach, we added fixed effects for specific states by adding dummies for each state, except one. Dr. Cunningham was kind enough to give us some of his time to discuss our approach and our interpretations, and we are very grateful for his help.

An alternative approach is outlined in “Difference-in-Differences with multiple time periods,” by Brantly Tawaya and Pedro Sant’Anna.^27^ In that article, they criticize certain elements of the two-way fixed effects regression approach and create their own statistical package to do the analysis that requires grouping first followed by regression.^28^ In parallel, another team of researchers – Borusyak et. al--released their similar model and code, as discussed in their article “Revisiting Event Study Designs: Robust and Efficient Estimation.”^29^ We group them together because they are similar in theory, although different in implementation. We used both methods to see if they produced consistent results.

### Method 3: A Machine Learning Approach Using a Causal Random Forest

As an alternative to a regression and the direct DID calculation, we used machine learning, specifically the causal random forest library because it is optimized to finding causal relationships, to see whether the independent variables can predict the dependent variable. If the approach works, we can identify which independent variables are the most important, and assess their impact that way. We wanted to see whether this approach produces results that are consistent with the regression approach.

At the end of the study, the causal random forest produced what we felt were anomalous results. They were inconsistent in both magnitude and direction from what the other models were telling us. As a consequence, we sat down with Teng Ye, an assistant professor from the University of Minnesota and an expert on causal random forest, to try to figure out why. Prof. Ye reviewed our data, and shared her opinion that in order to use a causal random forest and get a reliable result, we would need at least 3000-4000 data points. Our data frame has 665. She offered her opinion that our model was most likely overfitting. That resonated with us, particularly because each time we ran the model, we seemed to get a radically different result.

## X. Assumptions

### General Causal Inference Assumptions

The following are general assumptions needed for the causal inference to work reliably:

1. Conditional independence
2. Stable Unit Treatment Value Assumption (“SUTVA”)
3. Overlap Assumption
4. Exogeneity of covariates.^30^

Those assumptions are explained:

“The first and fourth assumptions, conditional independence and exogeneity of covariates, simply state that the treatment assignment is independent of the two potential outcomes and that the covariates are not affected by the treatment. The second assumption, Stable Unit Treatment Value Assumption (SUTVA), states that there is no interference or hidden variation between the treated and control observations. The third assumption, the overlap assumption, is that no subgroup is entirely located within either the treatment or control group. Consequently, the treatment probability has to be bounded away from zero and one, resulting in the propensity score.”^31^

Some of these assumptions may be problematic but we do not really know.

The first assumption gives us some concern, in that a state might adopt a CSA educational law in response to a rising problem of CSA, whereas other states with comparatively small CSA problems might not feel the need to adopt the law. This raises the problem of reverse causality, where CSA might cause the CSA educational laws.

While it is certainly possible, we would note that the rise of the CSA laws are also influenced by many other variables, including the presence of a vocal advocate. Enactment of laws also has much to do with the political environment in a given state. We also look at our data visualizations to see if the state level trends in the pre-adoption period were increasing or decreasing, and they appeared to be generally decreasing across all states. We also examined whether on average the per capita rate of CSA was greater in the adopting states, and found that indeed the per capita rate of CSA was slightly higher in states at the time at which they adopted the law, compared to the states that as of that juncture had not adopted the law.

However, the impact is not uniform across time, and on average is reasonably small. To interpret the chart above, it is important to also understand that there were differing numbers of states adopting the laws in different years, and actually a very large number of states adopted them in 2015 through 2019. In those years, the orange and blue lines are relatively close and indeed in the latter years orange goes below blue, meaning that the newly adopting states actually had less of a CSA problem than the never adopting states. On the whole, to us this means that if there is a reverse causation problem, it is not a large one.

Thus, we cannot rule out that the problem in some measure causes the solution. As a consequence, this did affect our choice of methodological tools. We did not treat the law as an instrumental variable because of this possibility, and instead chose the difference in difference methodology.

The second assumption – SUTVA – seems reasonable here because the legal process in one state should not have a big impact on what goes on in another state. However, we cannot rule out any impact. For example, if two states are side-by-side, and one has more lenient laws than the other, we can imagine a criminal deciding to cross the state boundary to commit his crime.

Further, as already explained, there were slight differences in the CSA educational laws. It is possible that those differences in the laws might impact the analysis.

With regard to the third assumption, our analysis does not include common subgroups across treatment and control states, so this should not be an issue.

We do not have a good way to test the independence of the covariates from the treatment, the fourth assumption. We are not particularly worried about the demographic covariates, as income and employment operate pretty independently of the legislative process around adopting CSA educational laws. But it is possible that states looked at the wide range of possible solutions to the CSA problem, and picked one or more than one and were consequently less likely to pick the other legislative solutions. At the same time, a single state could adopt any or all of the legislative solutions, and indeed there are several states that have more than one in place. We just do not have a good way of assessing the legislative process to determine whether adopting the various CSA legislative options is somehow not independent of the other legislative options.

Considering all of the assumptions together, we must confess that there is some possibility that our use of these causal inference tools is biased. We honestly do not know. But while these factors lead to potential inaccuracy, and suggest that we should not take specific numbers too seriously, we believe there is still some validity in the overall direction of our findings.

### DID Assumption--Parallel Trends

There are additional assumptions we need to make for the DID methodology to work. Specifically, we need to assume parallel trends.

We begin with the observation that there could be a variety of factors influencing the rate of CSA. As the research discussed above observes, such things as community morals impact the likelihood of CSA and those change over time and may change in different directions and in different ways in different states.

Beyond that, more practically, and more politically, we have seen the rise of survivors who act as advocates. According to the legal report that we use for the data in this project, “[c]hild sexual abuse prevention education laws in some states were enacted in memory of child victims, e.g.

Brooke Bennett in Vermont, Jeffrey Bell in Pennsylvania, or are named to honor adult survivors/advocates such as “Jenna’s Law” in Texas, “Jolene’s Law” in South Dakota, “Erin’s Law” and “Faith’s Law” in Illinois, “Tara’s Law” in Montana, “Bree’s Law” in Alaska. The impact these and many other survivors have had on legislation to prevent child sexual abuse cannot be overstated.” This localized activity almost certainly must impact the rate of CSA in those states, and not just adoption of the legislation.

Further, it is easy to believe here that there might be heterogeneous treatment effects that vary over time. We point out above that the laws actually differed slightly from state to state depending on such things as the grade levels affected and whether the laws were mandatory or voluntary. Apart from differences in the law themselves, there are undoubtedly differences in how they were implemented. As we noted above, we tried to quantify how vigorously schools across a given state implemented the law, but our social media strategy failed for lack of data. As a result, we have a question around how quickly and how extensively states implemented these laws.

As a result, our eyes are wide open when we look at the parallel trends assumption. We believe that there are probably some states that are quite different from others in their trends.

To examine this more closely, we created trend visualizations for all 50 states and studied them carefully. We then combined them to see how they related to each other. We made sure that we also adjusted for the conditions that we thought might impact the rate of CSA, including the population of the state (we would expect the number of cases to rise with larger population), but also economic factors such as unemployment and changes in income.

In the end, we also used software tools that are available to assess prior to the treatment whether the trends look similar. And the result we got there was that the trends generally look similar, but only to a confidence level of p = 0.17. Normally, as we observe later on, in scientific literature we look for a p <0.05. So a scientist might not view this as certain enough for a conclusion, but we nonetheless proceed with the analysis because we think there is value in it despite the uncertainties. Policy makers face the need to make decisions about what to do, and some information is better than no information, as long as it is understood in the proper context.

In doing this analysis, we were particularly guided by a recent World Bank blog post that in our opinion does a good job of laying out the best way to assess for parallel trends when there are multiple time periods.^32^ The blog post notes that “this assumption is that the untreated units provide the appropriate counterfactual of the trend that the treated units would have followed if they had not been treated – that is, that the two groups would have had parallel trends.”

We start by visualizing the trends together in one chart to see if they are roughly parallel.

As you can see, the trends across the 50 states look roughly the same. In this visualization, for simplicity, we called the 31 states that at some juncture adopted the laws the treatment states, and the controls are the never treated. We realize that in some of the algorithms below, treated states also can play the role of control states. But that issue aside, we want to simply depict the 50 states plus the District of Columbia, showing you the relevant trends.

The World Bank blog post also notes that “DiD will generally be more plausible if the treatment and control groups are similar in LEVELS to begin with, not just in TRENDS.” This visualization also illustrates the levels and shows that the majority of states are actually clustered below 1000 cases per year, with a sprinkling of states between 1000 and 4000 cases per year and then just a couple of outliers in the treatment category above 4000 cases. On the whole, this visualization suggests to us that the levels are comparable between the treated and the controls. The control and treatment states seem evenly distributed.

This visualization is the best we could come up with, but it is not perfect. For the control states, because there was no date of adoption to center it on, we simply started at 2009. So the dates do not really line up in that regard. That would be a problem, but for the fact that the control states are all pretty much smooth downward sloping trends. As you look at the trends, it really does not matter much exactly what year were talking about. They are without any abrupt changes and relatively constant in direction.

### Conditional Parallel Trend Assumption

While in one of our models we did not use controls, in most of them we did. So it is also useful to look at the data taking into account our controls. We visualize that in the following figure.

As already observed, we looked at all 50 states individually. The majority of them had declining trends prior to adoption of the law. That is one reason we are comforted, as we said above, that reverse causality is not too much of a confounding variable. Indeed, if you look at Alabama below, the decline after the law is actually less steep than the decline prior to the law. But the other thing we noted in examining all of the trends was that there were some states, as we mentioned above, where there was a very active advocate. Sometimes in such states there was an even steeper decline than we would have predicted, Illinois being an example.

In assessing parallel trends, we also thought it would be useful to look at the individual state trends in relation to the average trends among the never treated. The figure below shows the first six states to adopt the laws, and how those trends compared to the average never treated.

### Traditional Event Studies

We next performed event studies to see if there was any obvious change in the CSA case rate upon enactment of the laws. An event study takes all the different events that occurred at different times and lines them up at time zero to assess cumulatively whether the trend prior to the event changes after the event. When viewing an event study, it is important to mostly focus around the time periods closest to the treatment, as they are the most reliable. We did this twice, once using a traditional regression analysis and the other time using the Borusyak library.

In the first study, based on the regression analysis, the downward trend seems to continue without any impact from the treatment.

When we use the Borusyak library, the software shows the trend prior to and after the event, with a somewhat obvious decline after the event but the confidence levels continue to include zero as a possibility.

We also provide the Callaway event study below in Appendix B in the discussion of the context of discussing the substantiated CSA cases.

### Pretesting For Parallel Trends

The Callaway package provides a predesigned function for evaluating trends. As explained by Callaway, “although one cannot always test whether parallel trends itself holds, one can check if it holds in periods before treated units actually become treated. Importantly, this is just a pre-test; it is different from an actual test. Whether or not the parallel trends assumption holds in pre-treatment periods does not actually tell you if it holds in the current period (and this is when you need it to hold!).” That is why this is an assumption rather than a testable fact.

We ran the analysis twice, first without covariates and then with them. Without covariates, it appears that the model would suggest we have relatively high confidence that the parallel trends assumption is met prior to the event. Adding in the covariates though reduces our confidence to a p value of about 17%.

### Calculation Assumptions

The following summary adds three more specific assumptions for multiple time period DID as suggested in the “Introduction to DiD with Multiple Time Periods” by Callaway and Santa Anna published on June 29, 2021.^33^ It teases apart the parallel trends assumption to suggest that we ought to look at it both with only never treated units and then separately with not yet treated units.

- Staggered Treatment Adoption Assumption

“…Staggered treatment adoption implies that once a unit participates in the treatment, they remain treated. In other words, units do not “forget” about their treatment experience. …” Our study meets this test as once a state adopts the law, the law is in place for the remainder of the relevant time period.

- Parallel Trends Assumption based on never-treated units

“…This is a natural extension of the parallel trends assumption in the two periods and two groups case. It says that, in the absence of treatment, average untreated potential outcomes for the group first treated in time gg and for the “never treated” group would have followed parallel paths in all post-treatment periods t≥gt≥g. Note that the aforementioned parallel trend assumption rely on using the “never treated” units as comparison group for all “eventually treated” groups. This presumes that (i) a (large enough) “never-treated” group is available in the data, and (ii) these units are “similar enough” to the eventually treated units such that they can indeed be used as a valid comparison group. …” Again, our study meets this assumption and we designed our visualizations above to show this very point. We have 19 never treated states, and they represent a wide variety of states in terms of size and geographic region. The downward trends give us comfort that the parallel trends assumption is satisfied.

- Parallel Trends Assumption based on not-yet treated units

“…In plain English, this assumption states that one can use the not-yet-treated by time ss (s≥ts≥t) units as valid comparison groups when computing the average treatment effect for the group first treated in time gg. In general, this assumption uses more data when constructing comparison groups. However, as noted in Marcus and Sant’Anna,^34^ this assumption does restrict some pre-treatment trends across different groups. In other words, there is no free-lunch.” Our study design meets this assumption as well, as we make use of the not yet treated as controls. Many of the treatments occurred late in the relevant period, close to the end at 2019. So we used a considerable amount of data from those not yet treated states as controls. When you look at the chart, you will see that the pretreatment phase is reasonably parallel across all 50 states and the District of Columbia.

### Conclusion on parallel trends

On the whole, assumptions are just that; unproven assumptions. We have done our best to think them through, and above we reveal that there are places where the assumptions could be violated. On the whole, however, given the breadth of the data across all 50 states, and given what appears to us to be generally good evidence that these assumptions are reasonable, we believe that our conclusions are reasonably reliable. We would not claim that they are certain.

## XI. Analysis of the Evidence

Here we assess the evidence on the most important hypotheses. In the table below, we have reorganized the hypotheses slightly to tell a more coherent story.

**Table 1.**

|  | Scenario one | Scenario two | Scenario three |
| --- | --- | --- | --- |
| Reported CSA cases | **Decreases** because the reduction in actual crimes is bigger than the effect of causing previously unreported crimes to be reported | **Increases** because the effect of causing previously unreported crimes to be reported is bigger than the reduction in actual crimes | **Decreases** because the improvement in the quality of the reports offsets the fact that more people are reporting. |
| Substantiated CSA cases | **Decreases** because the law produces a decline in actual crimes. | **Increases** because the growth in reported crimes is bigger than the reduction in actual crime. | **Increases** because the growth in reported crimes is bigger than the reduction in actual crime. |

**Table 2:**
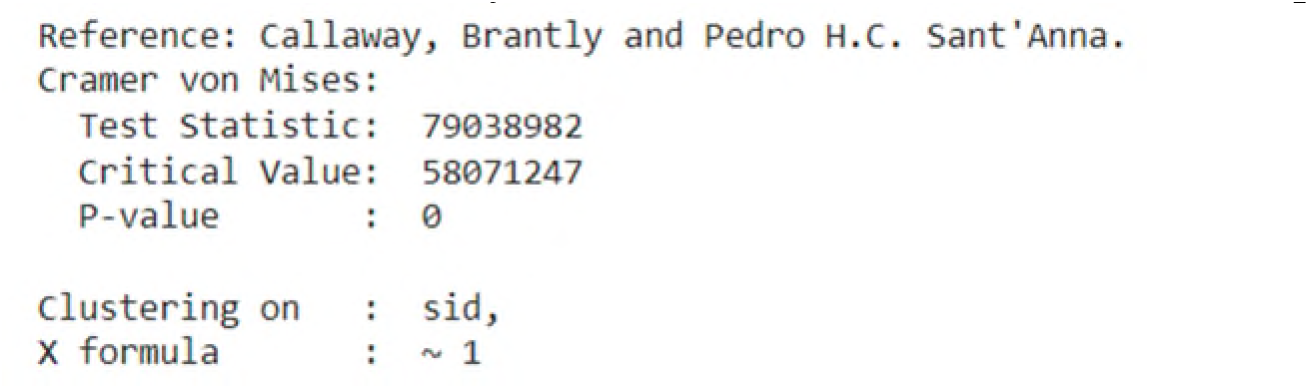
Statistical Analysis of Substantiated CSA Trends – Simple Model

**Table 3:**
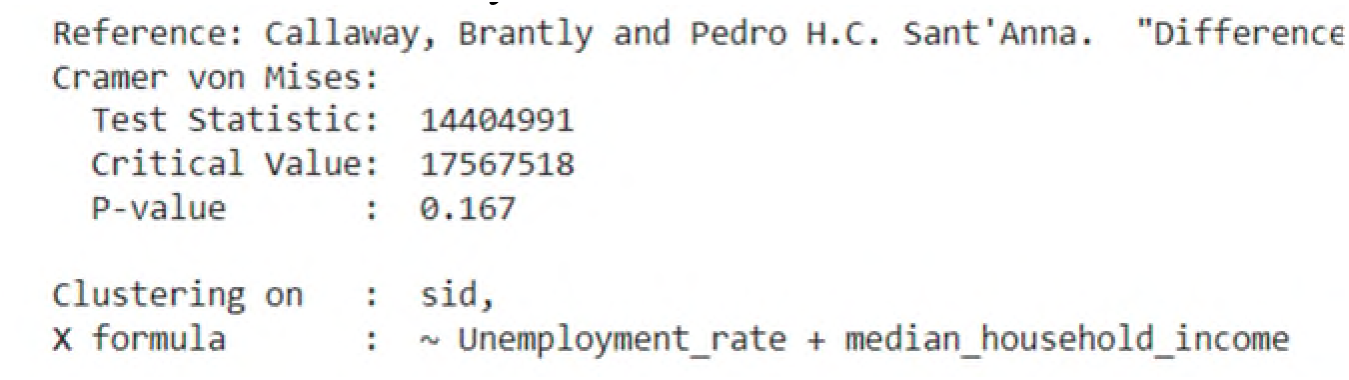
Statistical Analysis of Substantiated CSA Trends – With Economic Covariates

**Table 4:** Results from Primary Causation Tests

| Hypothesis | y Variable | Weighted Regression (Two Way Fixed Effects) | DID Groupings |  | Study Conclusion |
| --- | --- | --- | --- | --- | --- |
|  |  |  | Callaway and Sant'Anna | Borusyak, Jaravel and Speiss |  |
| NA, this row is covariates included in the models |  | Income, Unemp, Criminal, Poster, Screening, Task Force | Income, Unemp, Criminal | No covariates |  |
| 1, Reported CSA cases would change | Reported CSA Cases | 588** (299) | 539 (744) | 566 (621) | Reported cases went up, perhaps over 500 per state |
| 6, Ratio would go down (avg. 4.8) | Ratio reported/ substantiated CSA cases | 0.494** (0.233) | 0.030 (0.668) | 0.790 (0.368) | Ratio went up, perhaps by 0.5 |
| 2, Substantiated CSA cases would change | Substantiated CSA Cases | 21.1 (45.9) and -446*** (66.6)<br>1) | -212** (85) | -532** (194) | Substantiated CSA cases likely went down, but amount is uncertain, perhaps 200-500 |
| 7, Number of incompletely analyzed cases would remain the same | Incompletely Analyzed Reported CSA Cases | 0.329 (0.473) | -1.0228 (1.0214) | -0.989 (1.08) | No change |
| 4, Substantiated CA cases would change | Substantiated CA cases | -120 (662) | -2434 (2629) | -119 (2374) | No change—may be small drop but not statistically significant |

### Chart addendum

Standard errors are in parentheses. ** means statistically significant at p < 0.05 and *** means statistically significant at p < 0.01. The value in cell 1) was strongly negative and statistically significant when controlling for the economic variables as well as the other CSA laws, but when we added the linear trends the sign flipped to positive without statistical significance.

There are obviously differences as the reader goes across the chart looking at the results of the different methods we employed. The most obvious explanation for those differences is that each of the methodologies used a different number of covariates. We did not set out to do it this way, but it is the product of the methodologies themselves which do not all handle covariates equally well. It is also true that the methodologies themselves differ so we would not expect them to come up with identical results even with the same covariates.

We are convinced we see some causal effect in the three hypotheses highlighted in green. In those cases, we would reject our null hypothesis. Indeed, we also see a causal effect in hypothesis 6, but it is marked in red because we got the hypothesis exactly wrong. We predicted a decrease in the ratio, when it appears that there is a statistically significant increase in the ratio. So technically we would accept the null hypothesis, but we would also want to quickly reformulate our hypothesis to reflect what we learned about the ratio increasing.

In plain English, we state our conclusions in the final column. We think that the CSA educational laws caused the reported cases of CSA to increase, while also causing the substantiated CSA cases to decrease. We found no measurable impact on the substantiated cases of child abuse more broadly.

In the field of causation, it is all about assessing the totality of the evidence. Rarely is there a simple answer to the question of whether causation exists, but rather we consider the cumulated evidence to decide whether causation is likely, usually looking for a high level of confidence.

But it is important to keep in mind that this cannot be simply mathematically derived. We can do a statistical analysis through each one of our models. But as we are comparing our models and making judgments on which is more reliable, we must, for example, remember the assumptions we had to make and the strengths and weaknesses of each approach.

We want to emphasize that last point, that each way for assessing causation has strengths and weaknesses. The regression approach typically works well where the treatment effects are homogeneous, but breaks down if, for example, the later versus earlier comparisons add bias when there are dynamic treatment effects. Further, where the laws themselves are not completely uniform, it is reasonable to expect that we would see heterogeneous impacts among the various treatments. That, together with covariates that may change their impact over time, can bias the regression, as appears to have happened with the assessment of substantiated CSA cases. As we incrementally added control variables, we had strong negative numbers, but then when we added the linear trends, it flipped to a positive. It seems quite possible that is because of heterogeneous impacts.

The Callaway and Borusyak methodologies, which calculate DID without using regression (at least at first) by taking the first difference across time and then the second difference between states to arrive at a net effect, accommodates better any heterogeneous changes. Unfortunately, both of those implementations also have a weakness, in that the calculation becomes unwieldy as the user tries to add control variables. In our experience, going beyond just the economic control variables and one legal variable confounded the Callaway software and the Borusyak software did not even allow for covariates. That made it less reliable in our particular case because we felt we had several control variables that had significant impact.

We should also point out that reported CSA cases and substantiated CSA cases are fundamentally different variables and it is not unexpected that they might behave in different ways. The CSA educational law could influence the number of reported cases by primarily influencing the people who report such cases, while the number of substantiated cases is influenced by both the people who report and the caseworkers who do the substantiation. Our point is that focusing on fixed effects might make sense for one variable while not the other. Indeed, for example, the data seem to suggest that the number of substantiated CSA cases is more affected by heterogeneous effects than the reported cases.

In the end, although we would not rely entirely on any one model, we arrived at our conclusion on the basis of the consistency of the models and the fact that in each case, at least one model reported the results to be statistically significant. For reported CSA cases, the two-way fixed effects weighted regression across all four of our regression models predicted a statistically significant increase in the number of cases reported. The two DID direct calculations came up with a remarkably consistent impacts, although the standard errors they reported were larger. Given the consistency in the predicted effects and the statistical confidence of the two-way fixed effects regressions, we would be comfortable concluding that the number of reported CSA cases did indeed increase.

In the case of substantiated CSA cases, the two-way fixed effects weighted regression broke down because of the trending information. Over time, the heterogeneous impacts apparently biased that weighted regression model. But the two different direct DID calculations both indicated a decrease in the number of substantiated CSA cases in a statistically significant way. Given what appear to be changes over time, we find the DID calculations more reliable and sufficient to give us confidence in the outcome they report.

With regard to the ratio, we went with the two-way fixed effects again because across our four different regression models there was consistency. Further, at least in terms of magnitude of the change in the ratio, the Callaway and Borusyak models agreed, even though they put the standard error at a higher value. We were also comforted because this result would be consistent with an increase in reported CSA cases and a simultaneous decrease in substantiated CSA cases.

From a causation standpoint, we included the hypothesis about the incomplete handling of the reported cases to see whether the explanation for the ratio might be that something is limiting the capacity of the state departments of family services and their ability to handle an increase in reported CSA. But the numbers of incomplete cases did not seem to change in response to the laws, and therefore this does not seem to be a likely explanation for a decrease in the substantiated CSA cases.

In looking more broadly at all forms of child abuse, as opposed to simply CSA, while the law might have slightly reduced the number of substantiated CA cases, any impact was not statistically significant. Intuitively, it is not surprising that the impact on child abuse more generally would be smaller than on CSA.

We discuss our specific findings for each of the seven hypotheses in Appendix B. You will notice that we did more work to assess the causation of substantiated CSA cases than we did with the other y variables because substantiated CSA cases was our primary outcome. The other outcomes were simply necessary to get a fuller understanding of what was going on. We did not, for example, do event studies on the other y variables.

### Data exploration

Given the unexpected result in our sixth hypothesis, we were naturally interested in why the ratio went up instead of down. Without any particular hypothesis, we did some additional exploration we wanted to share so that future researchers could use it as perhaps a starting point. According to the code book that accompanies the data set, reported cases can be put into one of 10 different categories as follows:

1. Substantiated
2. Indicated or Reason to Suspect
3. Alternative Response-Victim
4. Alternative Response-Non-Victim
5. Unsubstantiated
6. Unsubstantiated-Intentionally False Report
7. Closed – No Finding
8. No Alleged Maltreatment 88. Other 99. Unknown Or Missing

There is also the category “Null” which means “Not Collected or Not Applicable.”

In our research, we focused on assessing the impact on number one. But since the results went in an unexpected direction, we wanted to explore the impact of these laws on the other possible answers.

There is a helpful discussion of what these other categories mean in the U.S. Department of Health and Human Services annual report on Child Maltreatment 2019 referenced above.^35^ The substantiated and unsubstantiated cases are the two most common answers, and they relate to whether the caseworker concluded that the allegation was supported or founded by facts and by state law.

While all states have those two possible answers, some states have adopted laws which would lead them to utilize other possible answers. For example, the category “indicated” in certain states means that the maltreatment could not be substantiated under state law, but there is reason to suspect that at least the child may have been maltreated or is at risk of maltreatment. The category of intentionally false is infrequently used, and its meaning is straightforward. The case worker believes that the report was maliciously made and not truthful. “Closed” with no finding means that the investigation by the child protective services organization could not be completed, often because they were unable to locate the alleged victim to interview. The no alleged maltreatment category is not as straightforward as it might seem. Apparently in a few states, there are laws requiring that the child protective services issue findings with respect to all children in the household when there has been an allegation involving one child, so the case worker might use this category to issue a conclusion for a child where there has been no even allegation of maltreatment. “Other” is what it typically means, the decision really does not fit into any of the designated categories.

The two alternative response categories are applicable to a few states that have adopted specific laws that direct the caseworkers not to investigate certain cases, typically because the cases are deemed low risk. This is a way of diverting low risk cases to be handled differently than a formal investigation.

Given those possible answers to the question, we wanted to explore using the same regression formulas what happened to the other variables. So we ran all the variables through the regressions, and most of them showed no statistically significant change, with three exceptions.

First, interestingly, the “other” category went down by about 140-290 cases, with a high statistical confidence of p = 0.01. Second, arguably even more interestingly, the category of “unsubstantiated” went strongly up by about 780-1160 cases, with equally high statistical confidence. Third, with equally strong confidence, the “unsubstantiated with intentionally false report” went down, but by a very small number, 3-5.

The other changes in the other variables were either not statistically significant or had some flip-flops of sign that indicated heterogeneous effects.

In addition to this set of regression analyses, we looked at the visualization of the data in all 50 states. There is great variation in which reporting options each state has based on their own state laws. There is also considerable variation across states with regard to even the major categories of substantiated and unsubstantiated.

### Summary of Movements in the Variables

The major movements in the variables seem to have gone something like this in numbers rounded to the 100s:

1. Reported up by about 500
2. Substantiated down by anywhere from 200 to 500
3. Other down by maybe 100 to 300
4. Unsubstantiated up by approximately 800 to 1200.

This appears to reconcile because if we have say 500 more reports, and let us say approximately 400 of the CSA cases are no longer in the substantiated category and 100 of the CSA cases are no longer in the “other” category, that leaves an unexplained gap of about 1000 reports which corresponds to the probable range of new reported cases.

The big question is why? Why did the education that followed these CSA educational laws produce these changes in the reported CSA cases? Unfortunately, the data do not tell us that. We would need data on what those educational programs were and how students and adults understood them in order to better understand how they might produce this effect. We will talk more about that below.

### Selection of Covariates

One of the great challenges of this research was obtaining data on relevant possible covariates. The research that we outlined above with regard to factors that may influence CSA rates include such things as individual factors (for example, the prevalence of disabilities), relationship factors (examples include drug abuse and marital dissolution among parents or other guardians), shifting community norms and shifting societal norms (including such areas as gender roles). There would appear to be much more going on that impacts the rate of CSA than we were able to capture in this study. Indeed, in the section below we identify future research, and expanding the covariates controlled while studying the mechanism of action would undoubtedly provide additional insights.

That said, we were able to control for certain features that did seem highly relevant from a public policy standpoint. For example, in each of the regressions, and most notably with regard to reported, substantiated and unsubstantiated CSA, the rate of unemployment played a significant role in predicting changes in our Y variable. On the other hand, to our surprise, median income seem to be much less relevant. The other laws that we control for that states have been adopting to address CSA also seemed to play a statistically significant role in affecting the rates of CSA. This is in particular screening, task force and poster laws. We were surprised to see such things as task force having an impact, because as laws go, such laws are rather benign. Perhaps, however, they are an indicator that states are focused on CSA and working on the issue.

While we were exploring the data, we made the decision to try to understand a little bit more about what each of the other legislative interventions contributed to impacting the rate of confirmed CSA cases beyond the information given us by the regressions. To do that, we utilized techniques for explanatory machine learning to produce the following insights from a random forest model.

First, we simply experimented with a random forest to assess feature importance, and learned as follows:

That was surprising to us, because the task force is the most benign of any legislative intervention. It basically accomplishes nothing other than saying that we will study the issue, and perhaps bringing media attention to the issue. Legally it really has almost no impact. Criminalization is a more dramatic step, although to be clear, CSA is always a crime, but this simply heightens the crime when committed by someone at a school. Post is simply the variable name we use for the legislation we are studying, the CSA educational laws.

We wanted to dig deeper, so we used the SHAP methodology to try to understand more specifically from our causal random forest model what was going on. As we explained above, our causal random forest model is unreliable because of the lack of adequate data to power it. But nonetheless, we thought it interesting.

As we expected, the demographic factors have the highest impact, with population being the greatest. That seems obvious, since the expected number of CSA cases is closely linked to the size of the population. Once we get past those demographic covariates, in this model it seems as though criminalization has a bigger impact on reducing the number of CSA cases modestly, while the task force approach in some cases produces the steepest decline in CSA cases. We do not have an explanation. Posters function like education but without the dynamic interactive component of teaching. Screening seems to have little impact. But again, we are not convinced that this causal random forest was producing accurate results so we would not put any weight on these results. We simply share them in the category of interesting.

## XII. Limitations

We face the following limitations in conducting this study, among others:

1. We are not measuring the thing we truly care about which is the total number of real CSA crimes committed in the United States. Instead, we are limited to only those CSA cases that get reported. We will never know the total number of actual cases that occur in a given year.
2. At best, we are able to estimate causal impact using a variety of techniques, each of which has its own strengths and weaknesses. The traditional use of weighted regression does not perform well when there are heterogeneous treatment impacts, and we believe that to be the case here because, among other things, the educational laws come in different flavors. The Callaway implementation of the DID calculation struggled to handle multiple control conditions. And the embodiment of the causal random forest that we used proved unreliable because of our small data set.
3. To use each of these estimating techniques, there are an associated list of assumptions that we need to make, most of which cannot be tested. We can make arguments as to why we think they might be valid, but we do not know for sure.
4. In this particular case, our treatment of interest, CSA educational laws, produces two opposing forces, one that would be expected to increase reporting and the other that would be expected to decrease cases to be reported. Given that we have no way to measure each of them separately, and we can only look at the net impact, we can only reach limited conclusions.
5. We have several control variables, and we could probably identify additional control variables.
6. Throughout this blog post we have been careful to say that we are focusing on the educational programs that resulted from the CSA educational laws. We worded that carefully for two reasons.

a. First, we are well aware that there were school programs before the laws were put in place. Thus, we are focused on the incremental increase in those school programs above what were previously utilized.
b. Second, we are also well aware that even after the law, many schools did not have the programs, either because the programs were only encouraged or because, if the programs were mandated, funds were not set aside to make those programs happen. It is quite common in legislatures to produce an unfunded mandate, a law that require something to be done but does not provide the resources to do it. These are simply part of our political process. Society does not always have the political will to allocate the money for things that we want done. Consequently, our analysis simply accepted all of this as a reality, and sought to measure the incremental improvement that resulted from adopting these laws. It may, however, be that the laws would have been more effective had they been appropriately funded. We cannot know.
7. We also were dealing with imperfect data in that a few states failed to submit all of the requested data over the 15 year period. That caused us to remove two states – Oregon and North Dakota – entirely, and fill in estimated values for single years for three more states. On the whole, that should not impact our calculation too much.

## XIII. Conclusion and Recommendation

Given that the system of reporting and substantiating CSA reports, and our hypotheses outlined above, here are the effects we found from states adopting CSA educational laws.

1. <u>The number of CSA reports increased as a result of the CSA educational laws</u>. This is perhaps not surprising, given that the purpose of the laws is to teach how to identify and report CSA, coupled with the historic underreporting of the offense. If the teaching is effective, we would expect people who had previously not understood what CSA was or how to report it to be able to do so in the future. Our regression analysis tells us that with p<0.05 confidence, depending on which other variables we control for, the CSA educational laws produced roughly 500 more reports per state per year on average. While we did not know what to expect, that result is not particularly surprising.
2. Our second finding is the most encouraging, in that it appears likely that <u>the number of substantiated cases of CSA resulting from the CSA educational laws went down</u>. Here we are relying on mostly the Callaway and Borusyak methods because the two-way fixed effects regression apparently was thrown off by the heterogeneous impact of the treatment over time. But both the Callaway and Borusyak methods suggest that this finding is statistically significant to a p<0.05.
3. What is *surprising* is the finding that <u>the quality of the CSA reports likely went down</u>. What we mean by that is that *proportionately fewer* of the reported CSA cases were later substantiated. We hypothesized the opposite. We theorized that teaching children and sometimes adults about how to accurately identify and report CSA would lead to a *higher* rate of substantiation of initial reports. But the results suggest the exact opposite effect. The ratio of reported to substantiated cases increased, meaning that there were more cases that were reported that were not then substantiated, after enactment of the CSA educational laws. How much? It appears that the laws resulted in an approximately 10% increase in that ratio. The average ratio over the 15 year period of the study was about 4.8, and the regression tells us that that ratio increased by about 0.46 as a result of the CSA educational laws. That is again with a p<0.05 confidence when using multivariate weighted regression.

The outcome was not consistent with our three theorized scenarios in section IV above. What we saw in the data was a fourth scenario where the two variables – reported and substantiated CSA – moved away from each other.

To move away from each other, this could potentially mean that the law was successful in deterring at least some CSA occurrences or helping children protect themselves, but at the same time caused more people to report CSA cases that could not be substantiated. The problem is, there is no way to tell from the data whether child were helped. Our further data exploration above suggests that in actuality the big trend is not so much that those two variables moved away from each other, but rather that many reports shifted to being unsubstantiated. We need to understand the reason behind the growth of the unsubstantiated variable before we can reach any conclusions.

Whether this over reporting of unsubstantiated cases is a problem may depend on who you ask. On the one hand, if you focus on victims, you may be pleased to hear that additional reports are being made, if you also believe where there is smoke there is fire. While it may be that an actual case of CSA has not yet not occurred, the report might prompt social workers to investigate enough such that CSA can be prevented or other needs the family has might be identified. For example, it may be that a potential future perpetrator is becoming overly friendly with a child with an ultimate intention of luring the child into a situation where the adult can commit sexual abuse, and by reporting that activity, even though it is not illegal, authorities can help prevent the actual crime from happening.

On the other hand, if you focus on people who might be wrongly accused simply because they are too friendly with children, this growth in reporting of unsubstantiated cases is concerning . Being accused of committing CSA is quite harmful. Our legal system and society are imperfect in many ways, and one way is that people who are inappropriately accused of a crime can nonetheless suffer even though the crime is never proven. People lose their jobs over such accusations, lose their relationships with friends and loved ones, and suffer many other losses. Unsubstantiated accusations of CSA in particular can be devastating because of the moral and social implications. If the CSA educational laws are actually increasing the number of unsubstantiated accusations, according to attorney thinking, that is a problem.

Fortunately, the past does not have to be the future. The tension we are describing above in the past forces us to weigh the need to protect children against the need to be fair to those accused. Different people will weigh those factors differently. But the future is wide open to us, and this tension is an artificial product of a system that does not have a method for reporting concerning but not illegal activity. If policymakers were to change the system going forward such that there is a separate means to report concerning but not illegal activity, that tension would mostly be gone.

We want to be clear that we do not have evidence that the overreporting is the fault of either the children or adults making the report, or the social worker recording the report. The problem more likely is that the current system has no alternative way of handling this information.

We should point out that this result also is not necessarily the fault of the statutes themselves, but rather may be more of a reflection on the way they were implemented. We talked throughout this paper of the problem of the unfunded mandate, i.e. a law that requires an action to be taken, but includes no corresponding appropriation of funds to do the necessary work. One take away from this is perhaps the need for a better, evidence driven approach to the educational content that is provided. In this improved program, for example, the content could focus on what exactly someone should do when they encounter grooming, instead of report it as CSA. Undoubtedly, this high-quality, standardized educational program would require funding to accomplish.

We wish to acknowledge the sheer complexity of the factors that go into deciding whether a given reported case of CSA ultimately is substantiated or unsubstantiated. Research in this area has identified a very wide range of factors that influence that decision by caseworkers, and those factors tend to get grouped into four categories:

- Case factors that include the situations events and circumstances related to the child and family.
- Decision-making factors that include caseworker characteristics such as training, experience, beliefs, their interactions with each other and their perceptions about the organization.
- Organizational factors including aspects of the child protective services work environment such as the structure of the agency, its resources tools and training, as well as workloads and supervision.
- External factors that include the state laws and other policies that govern the child protective services system.^36^

It is therefore well beyond the scope of our research to assess why the CSA educational laws might be influencing what appears to be a significant shift toward declaring more cases unsubstantiated.

Apart from CSA, we did not see evidence that the educational programs led to a reduction in the number of child abuse cases more broadly. We had theorized that some of the skills taught might help a child avoid abuse in other instances. However, in our analysis we saw no evidence of any statistically significant impact the CSA educational laws had on substantiated child abuse more broadly.

It is also important to put these findings in proper context. It would have been an unexpected pleasure if we had found that the CSA educational laws were by themselves a sort of silver bullet that substantially reduced the rate of CSA. Our clearest finding is that these CSA educational laws leave much of the problem left to be solved through other prevention strategies.

### Recommendations

We are concerned that the laws are significantly increasing the number of unsubstantiated reports of CSA, both through an increase in the number of reported cases but also through a decrease in the number of substantiated cases and cases placed in the “other” category. Our research does not lead us to conclude whether that is good or bad. In the best scenario, children are being helped, but at the expense of potentially hurting those who are innocently accused. In the worst scenario, children and the accused are hurt.

We therefore recommend that policymakers support additional research as we describe more in the next section to figure out why those changes are taking place. We think the most likely answer is found in the implementation of the laws. Somehow the education delivered caused more people to report alleged CSA, but more of those reports turned out to be unsubstantiated. Depending on the answer, the laws may be worthwhile to continue but with a modification to how the reporting is done, as more fully described in the following section. Such a modification would allow the benefit to children while avoiding the harm to wrongly accused adults.

## XIV. Future Research

As we explain in the analysis section, we conducted some additional data exploration and found that the additional cases were likely placed in the category of unsubstantiated. Future research could focus on the mechanism of action: specifically why are these CSA educational laws increasing the number of reported CSA cases and increasing the number of unsubstantiated CSA cases, at the partial expense of substantiated cases. We have no evidence on this mechanism question, only theories.

Going into this research, based on prior research discussed above, we believed that good education with regard to what constitutes CSA and how to report it should lead to more reports in general (as previously unrecognized incidents get reported) and specifically more reports that are can be substantiated (as we would have expected the reports to focus on incidents that constitute CSA as that term is defined in the law). There is considerable evidence as described above that it is possible to teach children and adults how to better identify CSA. So if this intuitive theory still has merit, there would appear to be other forces at work that lead on a net basis to a significant increase in the number of unsubstantiated cases, forces that outweigh those educational gains and more.

Following completion of our quantitative research, we spoke with experts and did further library research looking for an explanation of why unsubstantiated reports appear to have grown. Particularly the experts with whom we spoke suggested that much of the education may have sought to teach students and adults what to do if an adult gets inappropriately friendly with a child, often called grooming. If that is indeed what happened, then theoretically it would be possible that these adults and children would report that troublesome activity through the child abuse hotline in the state, thus making what we described above as a “referral” – the initial communication that ultimately might lead to a “report.” This theory could explain both the increase in the number of reports, but also the decline in the number of substantiated CSA cases. By getting a responsible adult involved through that reporting, that adult can work with the child or potentially the groomer to discourage the escalation of the relationship into CSA. If that is indeed what happened, the intervention might prevent the crime from occurring, thus leading to the decline in substantiated CSA cases.

In our research we identified no separate pathway presently for reporting grooming, since grooming is not illegal. As we previously observed, that was true throughout the time during which the data we are analyzing were created. ^37^ Going forward, obviously, policymakers could choose to create a separate reporting path. Our point is that if this theory turns out to be supported by the evidence, then going forward policymakers might choose to preserve the educational interventions because they are helping children but also implement a parallel reporting system to collect information on concerning but lawful activity that may need an intervention, to avoid implicating an adult who has not committed a crime.

But we cannot jump to the conclusion that that is in fact what happened. There are other possible explanations for why the unsubstantiated CSA cases grew. Here are just a couple of other theories that spring to mind.

Theory 2. The education was simply not effective at teaching what CSA is. However, the teaching was effective at generally raising the overall level of interest and awareness in the topic and in encouraging children and adults to report. The students and adults just did not learn well enough what to report. Under this theory, the reports went up because of the greater general interest and focus on the topic and encouragement to report. At the same time, it is possible that the CSA education was successful at deterring the crimes and teaching kids how to avoid being victims (even if they really did not learn how to more accurately identify the crimes). Thus, the number of actual crimes to be reported went down. Under this theory, if the number of actual crimes to be reported goes down, it is possible that that would produce the decrease in substantiated CSA cases at the same time stimulating what amount to be reports that cannot be substantiated.

Theory 3. We suppose it is possible that somehow this education has caused potential perpetrators to better understand the process, and to better hide the evidence against them such that the claims against them cannot be substantiated. If that is the case, these educational programs would be causing harm to children, and helping guilty perpetrators. This is pure speculation. We do not see evidence that this is what is going on, but we can also not rule it out.

Again, these are just theories developed after the research. We do not have any data to discern what actually caused the increase in the number of unsubstantiated cases.

We hope that researchers consider tackling why the number of unsubstantiated cases apparently rose in response to the CSA educational laws. It may not be possible at this juncture to go back in time to figure out why it happened as we do not believe there were any substantial data collected on how the educational programs were rolled out nationwide. There are, of course, the actual narrative reports and summaries of investigation that exist at the state level that were not reported up to the national database. Theoretically, if a researcher had access to those reports, they might provide some insight into why there was a growth in unsubstantiated cases. Given the highly sensitive nature that information, though, getting access to even a small portion of those state level reports would be quite difficult in terms of legal compliance and privacy protections.^38^

But there would be an opportunity going forward for researchers to collect information through a controlled trial on these educational programs in order to assess these theories prospectively. A controlled clinical trial would also offer the advantage of increasing confidence as a result of controlling for other possible covariates, something that we found challenging in the real-world data. As noted above, there are individual factors, family factors and community and social factors that we were not able to adequately control. However, a controlled trial here would be complex, large and expensive to implement. Alternatively, researchers could send questionnaires to caseworkers around the country to get a better understanding for why there exists this trend toward unsubstantiated in the face of these laws. Such a survey, however, would not be focused on the specific outcomes from the CSA educational laws, but rather trends more generally.

As we recited above in the section on prior learnings, the research already shows that it is quite possible to teach children and adults effective strategies. That is not the question. The question is assessing, among other things, what actually happens in the schools across the United States that perhaps do not have access to model educational programs on this topic, likely because they lack the financial resources to acquire them. We have not given much thought to how researchers might test theories three and four, which seem more difficult to evaluate, beyond the possibility of the survey mentioned above.

Education and child abuse experts could improve the education delivered through evidence-based, standardized content and techniques made available to schools nationwide. But if policymakers choose to go this direction, there should be a separate dedicated way of reporting lawful activity that is nonetheless troublesome, that improved content might need to include how to properly report such activity. This would relieve the tension that we described in our conclusions between helping children and avoiding needless injury to those inappropriately accused.

## Data Availability

All data produced in the present work are contained in the manuscript

https://github.com/BradleyMerrillT/ProjectPrevent

## Appendix A Detailed Analysis

Here we take a detailed dive into the evidence around our seven hypotheses.

All hypotheses are stated as alternative hypotheses. As you will see below, while we take the three basic analytic approaches to all seven hypotheses, in the case of hypotheses number two on substantiated CSA, we do a much deeper dive because that is the ultimate metric of interest.

- Hypothesis 1—Reported CSA Cases

### Educational programs on CSA that follow state enactment of laws requiring or encouraging such programs impacts the number of reported CSA cases

We reject the null hypothesis and conclude that the educational programs on CSA that follow state enactment of laws requiring or encouraging such programs do indeed impact the number of reported CSA cases. Here we rely primarily on the weighted regression analysis. Using the fourth model which includes all the covariates including the linear trends, we would estimate the reported number of CSA cases to increase by about 588 cases per state per year.

**Table 5:** Regression Coefficients for Reported CSA Cases

|  | *Dependent variable: rep_csa_cases* |  |  |  |
| --- | --- | --- | --- | --- |
|  | (1) | (2) | (3) | (4) |
| post[T.1.0] | 215.092<br>(240.218) | 534.188**<br>(240.398) | 627.703**<br>(253.501) | 588.189**<br>(299.256) |
| Unemployment_rate |  | 524.000***<br>(91.685) | 524.292***<br>(92.529) | 598.310***<br>(85.360) |
| median_household_income |  | -0.028<br>(0.027) | -0.030<br>(0.027) | -0.018<br>(0.027) |
| Screening[T.1.0] |  |  | 303.234<br>(498.294) | 737.381<br>(573.165) |
| Criminalization[T.1.0] |  |  | -45.767<br>(235.156) | 532.834**<br>(245.792) |
| Task_Force[T.1.0] |  |  | -405.297*<br>(242.569) | -2409.422***<br>(336.396) |
| Posters[T.1.0] |  |  | -42.205<br>(266.378) | -717.629**<br>(290.419) |
| Missouri_trend |  |  |  | 244.404**<br>(101.675) |
| Massachusetts_trend |  |  |  | 268.594***<br>(103.093) |
| Maryland_trend |  |  |  | 72.652<br>(108.631) |

**Table 6:**
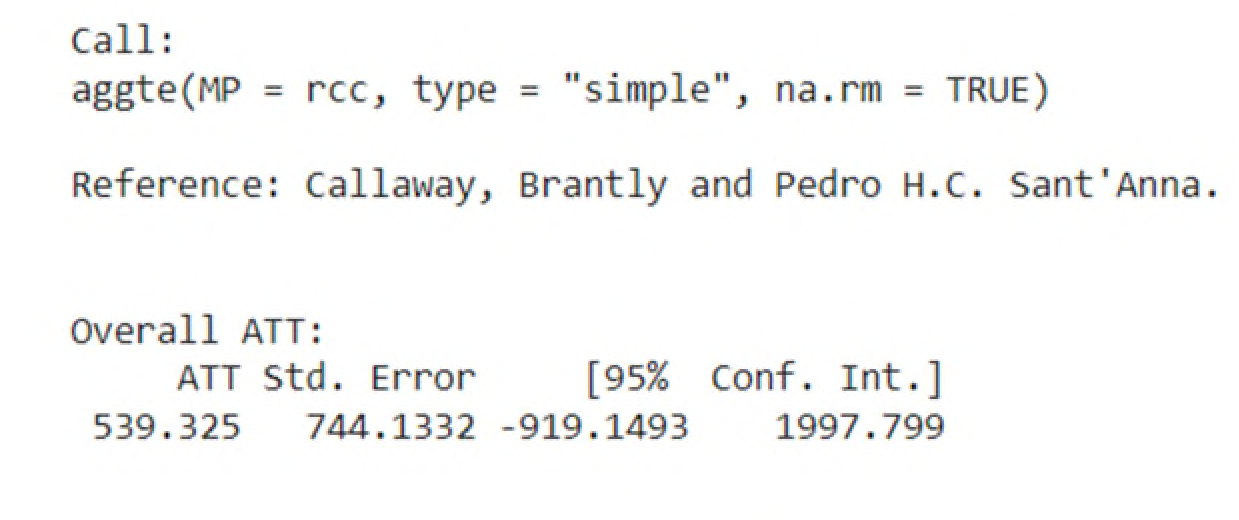
Callaway Calculations for Reported CSA Cases

**Table 7:**
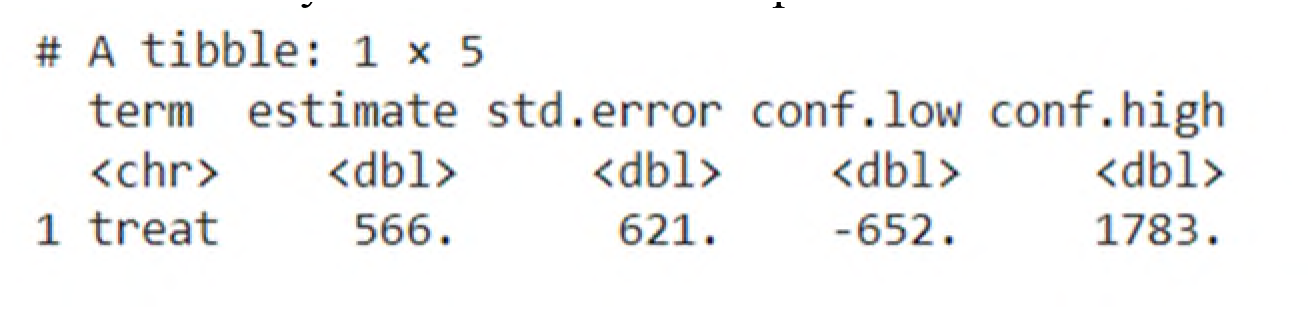
Borusyak Calculations for Reported CSA Cases

**Table 8:** Regression Coefficients for Substantiated CSA Cases

|  | *Dependent variable: sub_csa_cases* |  |  |  |
| --- | --- | --- | --- | --- |
|  | (1) | (2) | (3) | (4) |
| post[T.1.0] | -489.676***<br>(63.777) | -457.835***<br>(64.856) | -446.162***<br>(66.613) | 21.134<br>(45.853) |
| Unemployment_rate |  | 0.769<br>(24.735) | -7.480<br>(24.314) | -4.110<br>(13.079) |
| median_household_income |  | -0.027***<br>(0.007) | -0.023***<br>(0.007) | -0.002<br>(0.004) |
| Screening[T.1.0] |  |  | 18.435<br>(130.939) | -135.987<br>(87.822) |
| Criminalization[T.1.0] |  |  | -74.613<br>(61.793) | 211.159***<br>(37.661) |
| Task_Force[T.1.0] |  |  | 335.589***<br>(63.741) | -266.918***<br>(51.544) |
| Posters[T.1.0] |  |  | -234.748***<br>(69.997) | 2.402<br>(44.499) |
| Missouri_trend |  |  |  | -46.797***<br>(15.579) |
| Massachusetts_trend |  |  |  | -2.376<br>(15.796) |
| Maryland_trend |  |  |  | 12.396<br>(16.645) |

**Table 9:**
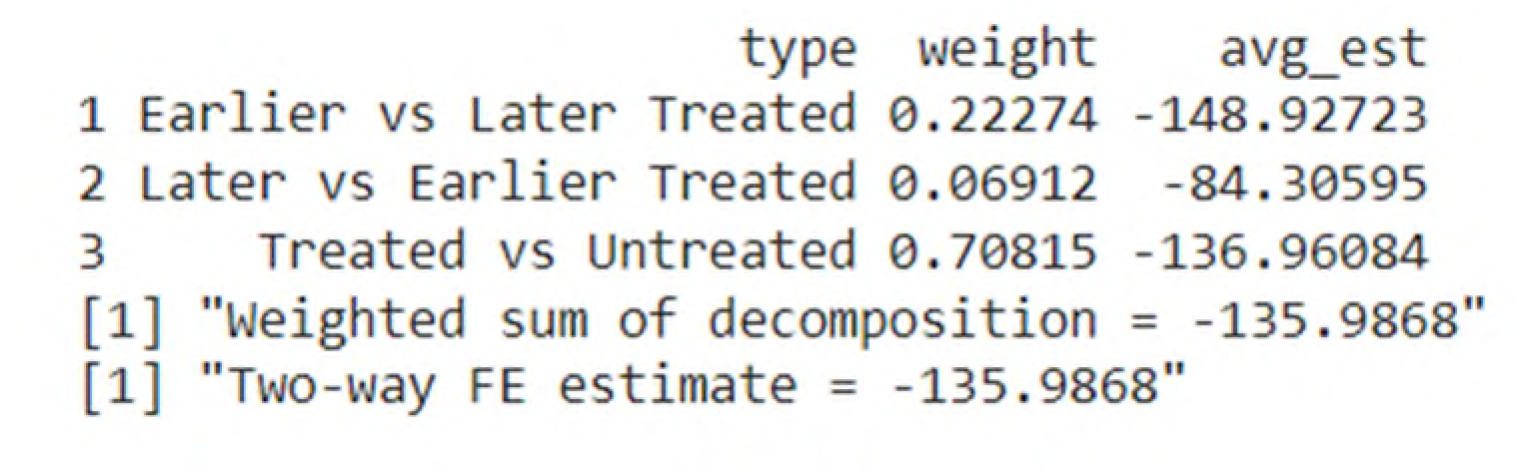
Goodman Bacon Decomposition for Substantiated CSA Cases

**Table 10:**
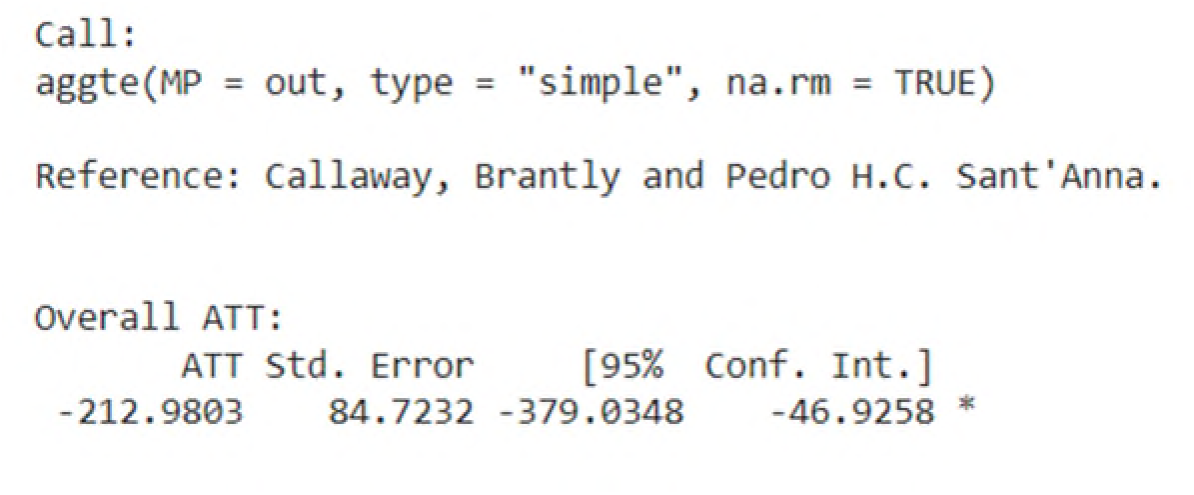
Callaway Calculations for Substantiated CSA Cases

**Table 11:**
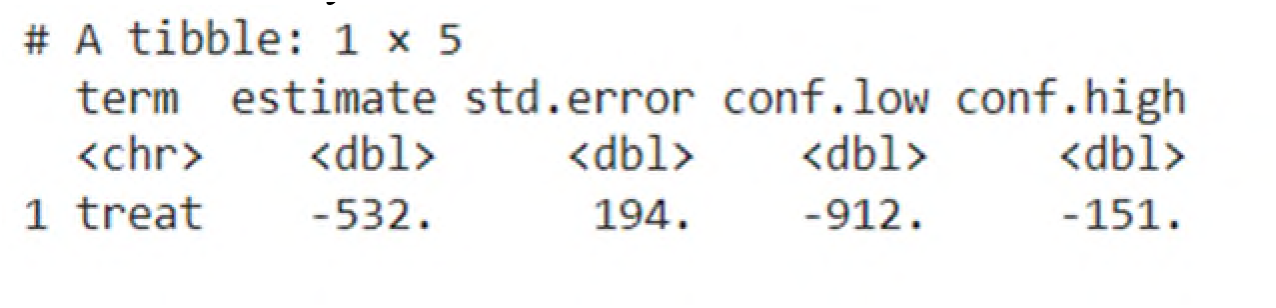
Borusyak Calculations for Substantiated CSA Cases

**Table 12:** Regression Coefficients for Substantiated CA Cases

|  | Dependent variable: sub_ca_cases |  |  |  |
| --- | --- | --- | --- | --- |
|  | (1) | (2) | (3) | (4) |
| post[T.1.0] | -759.651<br>(622.961) | -629.044<br>(640.003) | 45.334<br>(657.605) | -120.441<br>(662.267) |
| Unemployment_rate |  | 180.279<br>(244.091) | 62.760<br>(240.028) | -167.332<br>(188.905) |
| median_household_income |  | -0.027<br>(0.071) | 0.005<br>(0.070) | 0.007<br>(0.060) |
| Screening[T.1.0] |  |  | 3034.352**<br>(1292.620) | -18.561<br>(1268.439) |
| Criminalization[T.1.0] |  |  | -1079.330*<br>(610.016) | -771.549<br>(543.948) |
| Task_Force[T.1.0] |  |  | 2165.724***<br>(629.245) | -2253.008***<br>(744.459) |
| Posters[T.1.0] |  |  | -3176.324***<br>(691.009) | -1480.034**<br>(642.709) |
| Missouri_trend |  |  |  | 176.917<br>(225.010) |
| Massachusetts_trend |  |  |  | -448.483**<br>(228.150) |

**Table 13:**
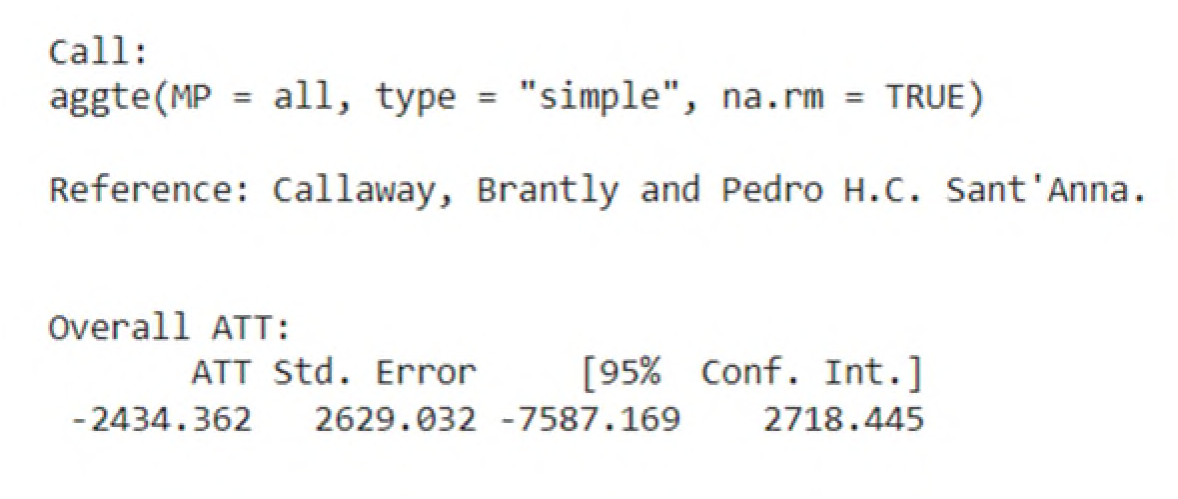
Callaway Calculations for Substantiated CA Cases

**Table 14:**
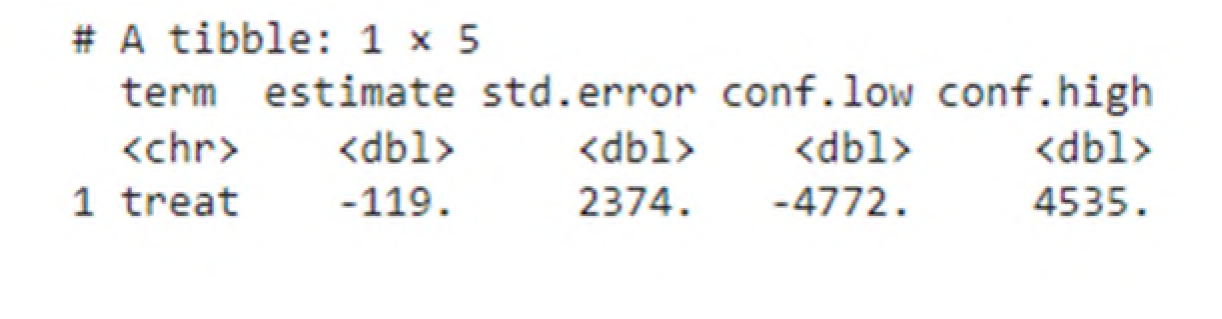
Borusyak Calculations for Substantiated CA Cases

**Table 15:**
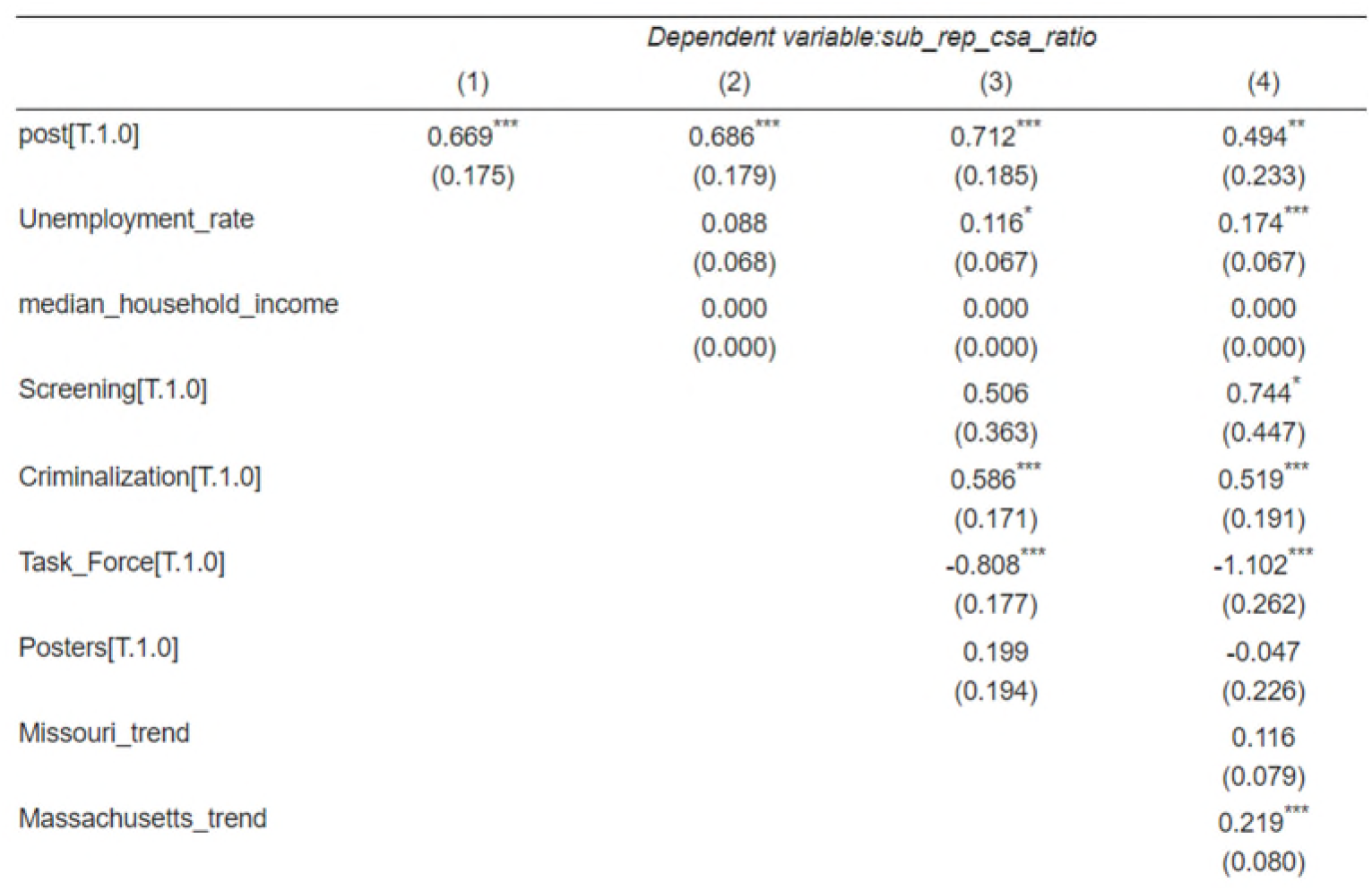
Regression Coefficients for Ratio of Reported to Substantiated CSA Cases

|  | Dependent variable: sub_rep_csa_ratio |  |  |  |
| --- | --- | --- | --- | --- |
|  | (1) | (2) | (3) | (4) |
| post[T.1.0] | 0.669***<br>(0.175) | 0.686***<br>(0.179) | 0.712***<br>(0.185) | 0.494**<br>(0.233) |
| Unemployment_rate |  | 0.088<br>(0.068) | 0.116*<br>(0.067) | 0.174***<br>(0.067) |
| median_household_income |  | 0.000<br>(0.000) | 0.000<br>(0.000) | 0.000<br>(0.000) |
| Screening[T.1.0] |  |  | 0.506<br>(0.363) | 0.744*<br>(0.447) |
| Criminalization[T.1.0] |  |  | 0.586***<br>(0.171) | 0.519***<br>(0.191) |
| Task_Force[T.1.0] |  |  | -0.808***<br>(0.177) | -1.102***<br>(0.262) |
| Posters[T.1.0] |  |  | 0.199<br>(0.194) | -0.047<br>(0.226) |
| Missouri_trend |  |  |  | 0.116<br>(0.079) |
| Massachusetts_trend |  |  |  | 0.219***<br>(0.080) |

**Table 16:**
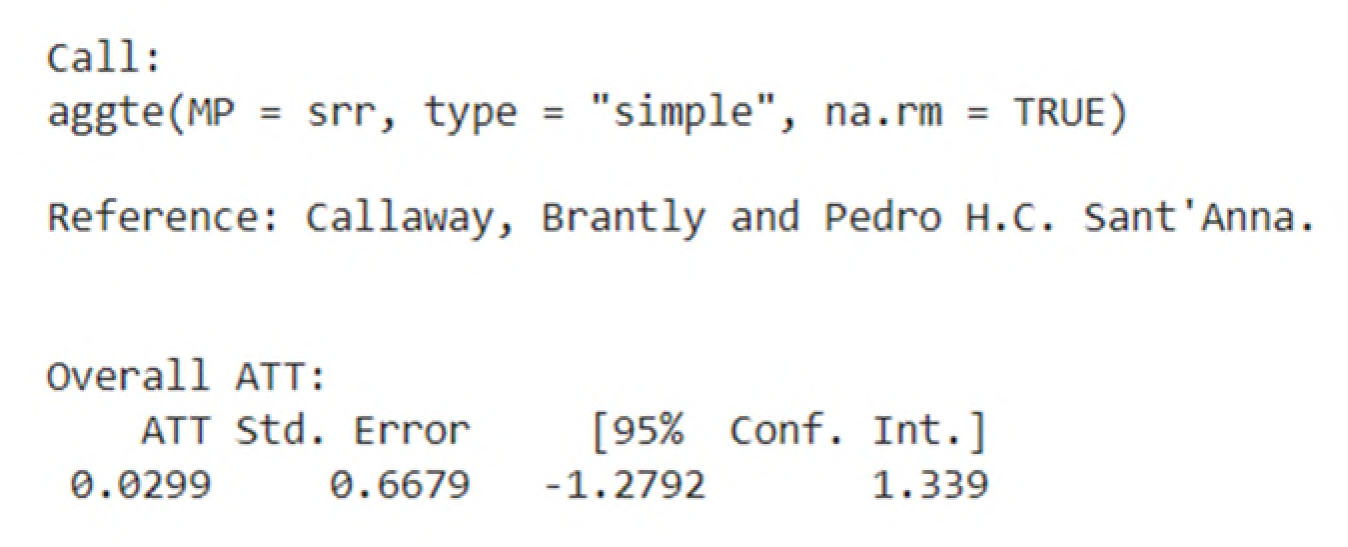
Callaway Calculations for Ratio of Reported to Substantiated CSA Cases

**Table 17:**
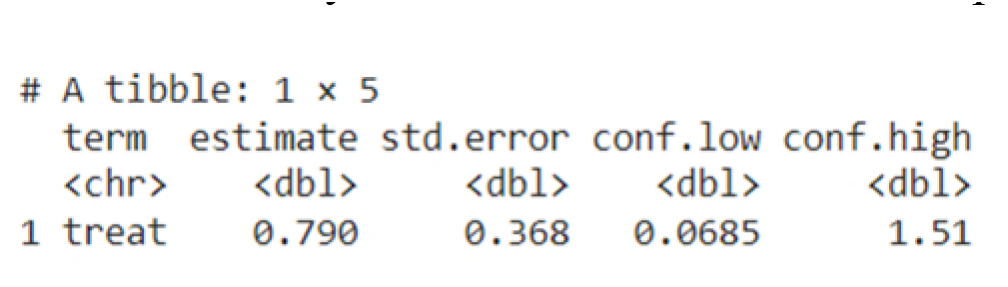
Borusyak Calculations for Ratio of Reported to Substantiated CSA Cases

**Table 18:** Regression Coefficients for Incomplete Substantiated CSA Cases

|  | *Dependent variable: rep_csa_inc* |  |  |  |
| --- | --- | --- | --- | --- |
|  | (1) | (2) | (3) | (4) |
| post[T.1.0] | -0.115<br>(0.416) | -0.205<br>(0.427) | -0.162<br>(0.449) | 0.329<br>(0.473) |
| Unemployment_rate |  | -0.251<br>(0.163) | -0.254<br>(0.164) | -0.162<br>(0.135) |
| median_household_income |  | -0.000<br>(0.000) | -0.000<br>(0.000) | 0.000<br>(0.000) |
| Screening[T.1.0] |  |  | 1.175<br>(0.882) | -1.377<br>(0.906) |
| Criminalization[T.1.0] |  |  | 0.613<br>(0.416) | -0.407<br>(0.388) |
| Task_Force[T.1.0] |  |  | 0.592<br>(0.429) | 0.647<br>(0.531) |
| Posters[T.1.0] |  |  | -0.715<br>(0.472) | 0.156<br>(0.459) |
| Missouri_trend |  |  |  | -0.032<br>(0.161) |
| Massachusetts_trend |  |  |  | 0.524***<br>(0.163) |

**Table 19:**
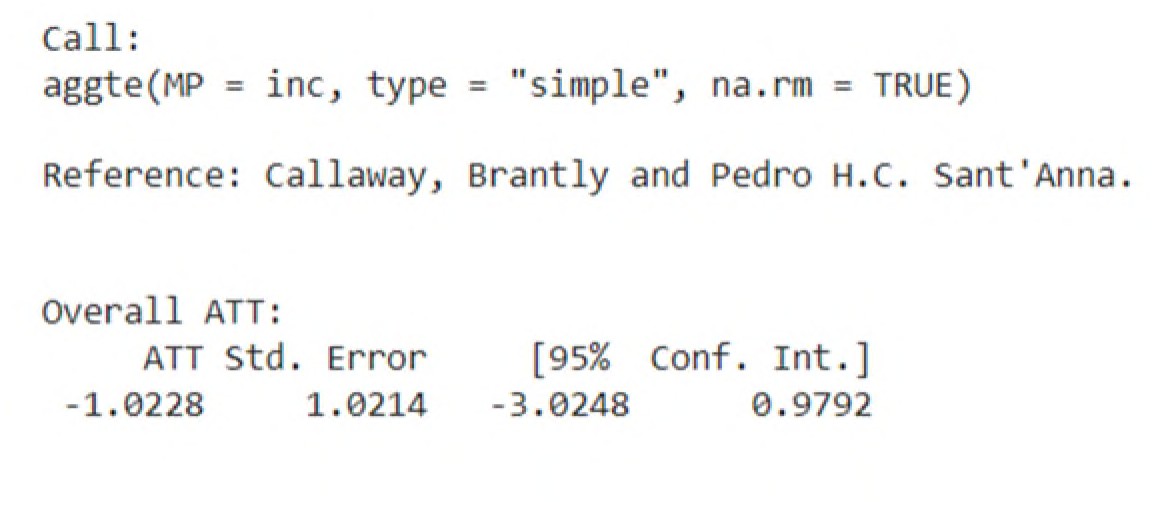
Callaway Calculations for Incomplete Substantiated CSA Cases

**Table 20:**
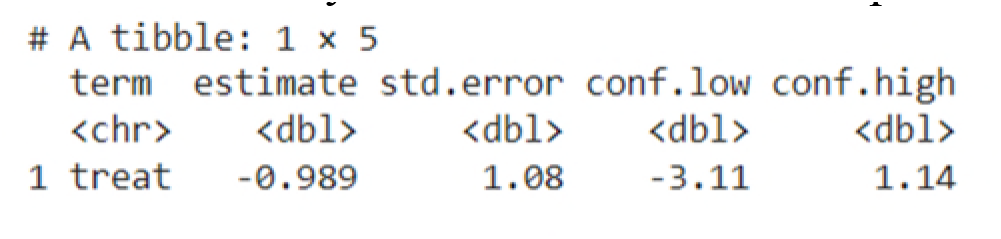
Borusyak Calculations for Incomplete Substantiated CSA Cases

**Table 21:**
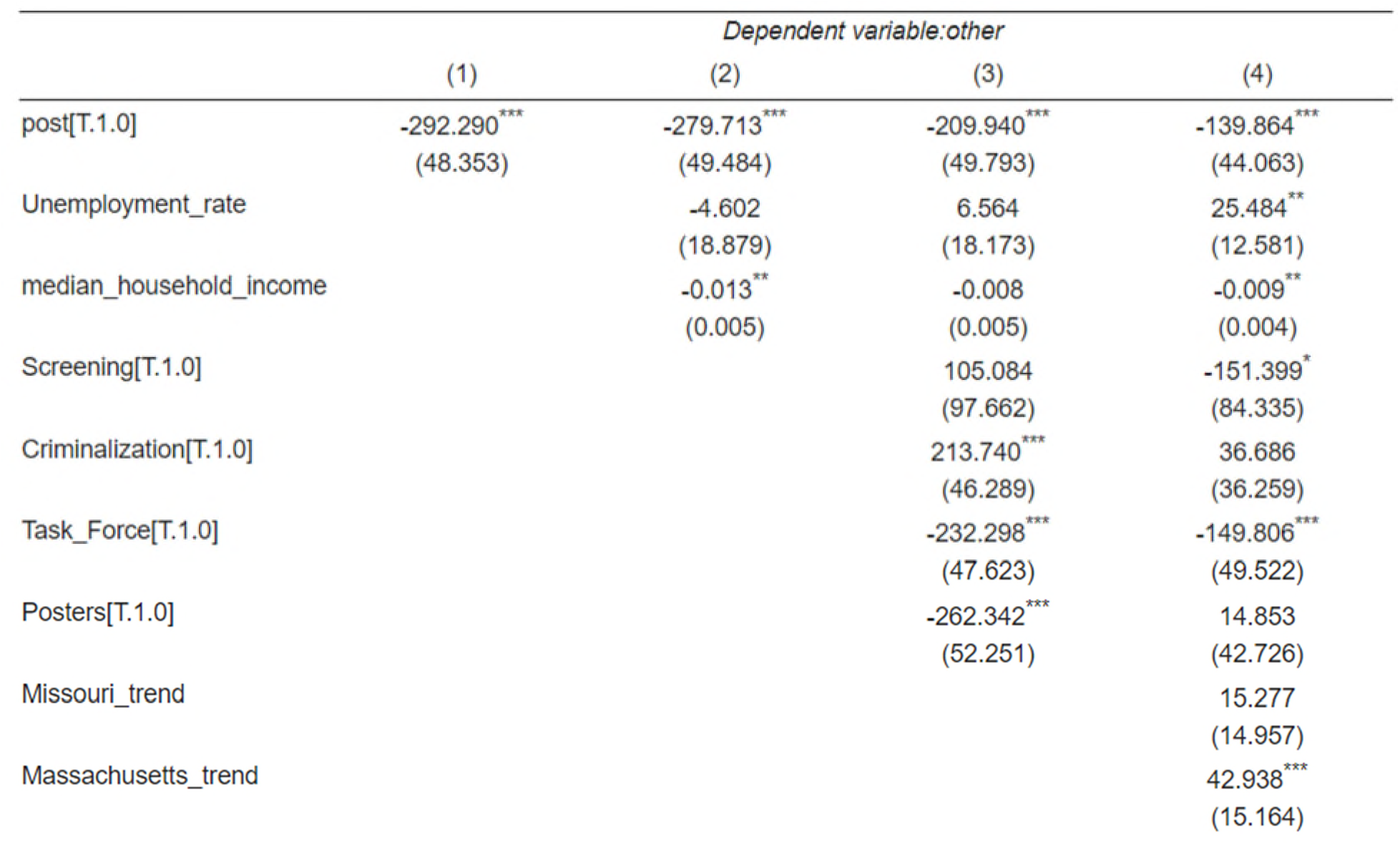

|  | *Dependent variable: other* |  |  |  |
| --- | --- | --- | --- | --- |
|  | (1) | (2) | (3) | (4) |
| post[T.1.0] | -292.290<sup>***</sup><br>(48.353) | -279.713<sup>***</sup><br>(49.484) | -209.940<sup>***</sup><br>(49.793) | -139.864<sup>***</sup><br>(44.063) |
| Unemployment_rate |  | -4.602<br>(18.879) | 6.564<br>(18.173) | 25.484<sup>**</sup><br>(12.581) |
| median_household_income |  | -0.013<sup>**</sup><br>(0.005) | -0.008<br>(0.005) | -0.009<sup>**</sup><br>(0.004) |
| Screening[T.1.0] |  |  | 105.084<br>(97.662) | -151.399<sup>*</sup><br>(84.335) |
| Criminalization[T.1.0] |  |  | 213.740<sup>***</sup><br>(46.289) | 36.686<br>(36.259) |
| Task_Force[T.1.0] |  |  | -232.298<sup>***</sup><br>(47.623) | -149.806<sup>***</sup><br>(49.522) |
| Posters[T.1.0] |  |  | -262.342<sup>***</sup><br>(52.251) | 14.853<br>(42.726) |
| Missouri_trend |  |  |  | 15.277<br>(14.957) |
| Massachusetts_trend |  |  |  | 42.938<sup>***</sup><br>(15.164) |

**Table 22:**

|  | *Dependent variable:un* |  |  |  |
| --- | --- | --- | --- | --- |
|  | (1) | (2) | (3) | (4) |
| post[T.1.0] | 783.349<sup>***</sup><br>(225.220) | 1077.069<sup>***</sup><br>(224.430) | 1155.729<sup>***</sup><br>(235.146) | 813.917<sup>***</sup><br>(258.879) |
| Unemployment_rate |  | 548.712<sup>***</sup><br>(85.625) | 550.833<sup>***</sup><br>(85.824) | 612.435<sup>***</sup><br>(73.919) |
| median_household_income |  | 0.002<br>(0.025) | -0.007<br>(0.025) | -0.005<br>(0.024) |
| Screening[T.1.0] |  |  | 165.960<br>(461.210) | 1063.775<sup>**</sup><br>(495.482) |
| Criminalization[T.1.0] |  |  | -183.329<br>(218.599) | 319.378<br>(213.029) |
| Task_Force[T.1.0] |  |  | -728.956<sup>***</sup><br>(224.900) | -1824.595<sup>***</sup><br>(290.949) |
| Posters[T.1.0] |  |  | 299.433<br>(246.754) | -825.294<sup>***</sup><br>(251.021) |
| Missouri_trend |  |  |  | 278.270<sup>***</sup><br>(87.878) |
| Massachusetts_trend |  |  |  | 221.529<sup>**</sup><br>(89.093) |

**Table 23:**

|  | *Dependent variable: udifr* |  |  |  |
| --- | --- | --- | --- | --- |
|  | (1) | (2) | (3) | (4) |
| post[T.1.0] | -4.377***<br>(1.283) | -5.105***<br>(1.311) | -4.327***<br>(1.378) | -3.583***<br>(1.382) |
| Unemployment_rate |  | -1.381***<br>(0.500) | -1.466***<br>(0.503) | -0.546<br>(0.395) |
| median_household_income |  | -0.000<br>(0.000) | -0.000<br>(0.000) | 0.000<br>(0.000) |
| Screening[T.1.0] |  |  | 2.510<br>(2.704) | 0.302<br>(2.646) |
| Criminalization[T.1.0] |  |  | -2.214*<br>(1.281) | -0.108<br>(1.137) |
| Task_Force[T.1.0] |  |  | -2.585*<br>(1.318) | -0.520<br>(1.553) |
| Posters[T.1.0] |  |  | 0.259<br>(1.446) | 2.578*<br>(1.340) |
| Missouri_trend |  |  |  | -0.065<br>(0.469) |
| Massachusetts_trend |  |  |  | 0.161<br>(0.476) |

As you can see in the third regression model, taking into account the other laws actually increases the expected impact of the educational laws. Further, the regression analysis stays reasonably consistent even when the linear trends are introduced, suggesting that we can rely on it.

The Callaway model predicts about the same, but that is without taking into account several of the legal covariates. As already mentioned, the model fails to include many of the covariates that we use in the regression analysis

The Borusyak model takes into account none of the covariates, but still comes up with the same general magnitude result. Like Callaway, it also has a higher standard error.

- Hypothesis 2— Substantiated CSA Cases

### Educational programs on CSA that follow state enactment of laws requiring or encouraging such programs impacts the number of substantiated CSA cases

We reject the null hypothesis, meaning that we conclude that the laws do impact the number of substantiated CSA cases.

The weighted regression shows signs that the Y variable here is heterogeneous among the states, and therefore not reliable. In our four different weighted regressions models, the first three were all strongly negative, while the fourth was positive after introducing the linear trends. That switch of signs when linear trends are introduced is indicative of heterogeneous effects.

Goodman-Bacon proposes a series of diagnostic tests to examine the robustness of the two-way fixed effects DID estimate. He is concerned that late adopters are used as a control group for early adopters, but early adopters are also a control group for late adopters. He argues that heterogeneous treatment effects across both groups in time may lead to severe bias. Getting the right estimate depends on getting the weights correct. However, units that are treated either early or late receive very little weight in the two-way fixed effects estimation. Mathematically, the DID regression gives the greatest weight to groups whose treatment periods are 50% of the sample period apart.

He recommends decomposing the individual 2×2 DID weights, and then using a more effective balancing test for staggered DID.^39^ Specifically, Goodman-Bacon proposes a single t-test of the reweighted balance against the null of no imbalance. Note how much difference the later versus earlier treated result is than the others.

Notice how the later versus earlier treated gets very little weight.

It is also helpful to test the stability of the DID coefficients by plotting each 2×2 DID against its weight. We can calculate a variety of different conditional expectations over the subgroups, including the average effect and total weights for treated/untreated comparisons, and late/early treatment and early/late treatment comparisons. Adding the weights on the timing-based coefficients from the decomposition show how much of the estimate comes from timing variation.

**Figure 23:**
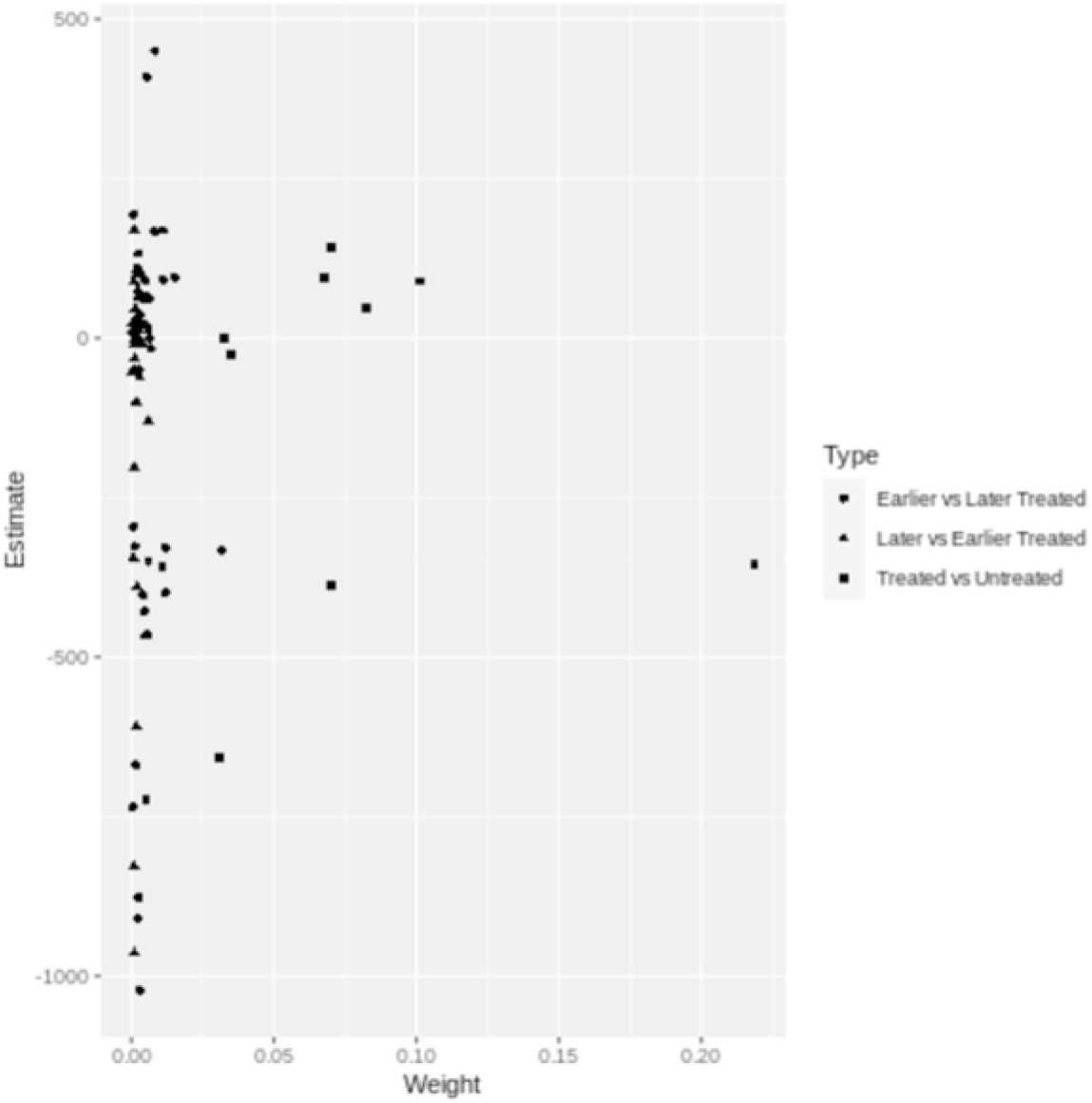
Goodman Bacon Decomposition Weights for Substantiated CSA Cases

This chart amplifies the statistics provided above, and shows that the later versus earlier treated get very little weight, but are clustered to the negative.

### Callaway Analysis

As already indicated, the Callaway software performs the basic DID calculation, but struggles sometimes with covariates. The simple output includes the ATT, and the associated standard error. Here it suggests a negative ATT for the substantiated CSA cases with statistical significance noted.

The Borusyak likewise predicts a negative treatment effect, and also concludes that it is statistically significant.

The concurrence of those two DID calculations both finding a negative result in both finding it to be statistically significant gives us confidence to reject the null hypothesis. The results are roughly similar to the first three multivariate regression models as well.

The Callaway software offers a feature for group analysis, grouping the treatments by the year of the treatment. It then calculates the ATT for each particular group.

- Hypothesis 3—All Child Abuse

### Educational programs on CSA that follow state enactment of laws requiring or encouraging such programs impacts number of substantiated <u>child abuse cases</u>, as opposed to simply CSA cases

Again we fail to reject the null hypothesis. The CSA educational laws do not appear to impact the rate of child abuse more generally. The theory was that the skills kids learn in the context of identifying and reporting CSA might transfer over to other abuse cases.

The regression coefficients alternated between positive and negative, and ended up much smaller than the standard error. It appears to be another dry hole.

The Callaway and Borusyak analyses likewise produced a wide standard error that easily included zero.

There are probably at least two possible explanations for why we found nothing in the area of impact on child abuse generally. The first is that the changes we found in the context of CSA were relatively modest, and we would expect any secondary improvement in child abuse generally to be smaller/weaker than the effect we observed for CSA. So it may be that the impact was simply so small that we did not see anything statistically significant.

Further, it is possible that the theory is simply wrong, that the self-protection skills learned in the context of CSA are indeed specific to CSA, and do not carry over to any substantial degree to child abuse more generally. The data really do not give us an insight into which of those two theories is the more likely explanation for why we did not find anything.

- Hypothesis 4—Ratio of Reported to Substantiated

### Educational programs on CSA that follow state enactment of laws requiring or encouraging such programs decrease the ratio of reported CSA cases to substantiated CSA cases

While we went out on a limb here and hypothesized that the educational programs would decrease the ratio of reported to substantiated CSA cases. Our theory seemed intuitively safe to us. Turns out we were on the wrong limb. Indeed, not only does it appear that we failed to reject the null hypothesis meaning that the law likely has *no such decrease*, it actually appears as though the law *increased* the ratio of reported to substantiated CSA cases. The exact opposite of our prediction. We were way off. It appears that by the time we factor in all of the covariates including the linear trends, the ratio of reported to substantiated cases goes up by almost 0.5. Given that the ratio is in the vicinity of 5.0, that means an almost 10% increase in the rate of reported to substantiated cases.

That surprised us, especially since previously we found a statistically significant increase in reported cases. So reported cases likely went up. The ratio of reported to substantiated likely went up. The number of substantiated likely went down.

Unfortunately, our analysis cannot take us much farther than that to discern exactly what caused the rate of substantiated to go down in the face of an increase in reported cases. We can come up with theories, but the data do not give us evidence to test those theories.

We did, of course, want to look at the other methodologies to see if they produced any different assessment with regard to the ratio. With regard to the Callaway calculation, it actually suggested a very small increase, but with a large standard error. The confidence interval is very wide and almost centered on zero.

The Borusyak is actually much closer to the regression estimate than it is to the Callaway estimate. It has a standard error half the size of Callaway, and the result comes very close to being statistically significant like the regression estimate.

Given the fairly close alignment between the regression estimate and the Borusyak Calculations, both of which are statistically significant or extremely close to statistically significant, we feel comfortable concluding that the ratio went up. That also aligns with the findings with regard to the direction of the reported and substantiated cases.

- Hypothesis 5—Incomplete Investigations

### Educational programs on CSA that follow state enactment of laws requiring or encouraging such programs do not affect the number of incomplete investigations of reported CSA cases

The challenge in demonstrating causation is that a researcher really needs to try to rule out all possible alternative explanations. For example, as the number of reported cases increases, presumably more social workers would be needed in order to handle the workload and substantiate the reports. Theoretically, it would be possible that perhaps the reason that more cases were not substantiated has nothing to do with the law, but instead the resources needed to hire the social workers who would substantiate the cases.

While we do not have data on the number of social workers, we do have data on the number of reports simply left open. Leaving a report open would be some evidence that the state departments of child services did not have the resources, or perhaps the cooperation of the reporter, to close the matter as either substantiated or not. We looked for any change in the number of open cases, and but found no relationship between enactment of the laws and any change in the number of cases unresolved as substantiated or not.

For the regression analysis, like a couple of examples above, the sign flips when we add the trends covariates. The heterogeneity makes the regression inconclusive.

The Callaway, with its shortcomings obviously still, predicts a small decrease in the number of incomplete actions on the reports, but has a standard error bigger than the coefficient. Zero is easily within the confidence interval.

The same is true of the Borusyak Calculations.

It seems that the methodologies all point in the direction of inconclusive, which generally means that if there is a causal impact, it is small. That suggests to us that resource constraints such as on the number of social workers is unlikely to explain why, in response to the CSA educational laws, the number of substantiated CSA cases may have dropped.

### Data exploration

Given the unexpected result in our sixth hypothesis, we were naturally interested in why the ratio went up instead of down. As explained in the analysis section, we did some exploration with regard to the impact on the other variables.

First, interestingly, the other category went down, with a high statistical confidence of p = 0.01.

Second, arguably even more interestingly, the category of unsubstantiated went strongly up with equally high statistical confidence.

Third, with equally strong confidence, the unsubstantiated with intentionally false report went down, but by a very small number.

## Footnotes

1 Preventing Child Sexual Abuse |Violence Prevention|Injury Center|CDC. (2021, April 30). Retrieved August 16, 2021, from https://www.cdc.gov/violenceprevention/childsexualabuse/fastfact.html

2 A Call to Action for Policymakers and Advocates: Child Sexual Abuse Prevention Legislation in the States. (2021, June). Retrieved August 16, 2021, from https://www.enoughabuse.org/images/Legislation/A_Call_to_Action-June2021.pdf

3 Preventing Child Sexual Abuse |Violence Prevention|Injury Center|CDC. (2021, April 30). Retrieved August 16, 2021, from https://www.cdc.gov/violenceprevention/childsexualabuse/fastfact.html (This section largely flows from that webpage, because we are conducting our research in response to issues CDC raised.)

4 Chapter 2, Child Maltreatment 2019, Report by the United States Department of Health and Human Services. January 14, 2021 accessed December 17, 2021 at https://www.acf.hhs.gov/cb/report/child-maltreatment-2019

5 Here, for example, is a description of the process in California. What Happens After a Child Sexual Abuse Report Is Filed? (2020, May 18). Retrieved November 17, 2021, from https://www.manlystewart.com/what-happens-after-a-child-sexual-abuse-report-is-filed/

6 Child Sexual Abuse Prevention Darkness to Light. (2021, October 01). Retrieved from http://www.d2l.org/; http://www.d2l.org/wp-content/uploads/2017/01/Statistics_6_Reporting.pdf

7 Indeed, here is a list of the top 10 reasons people do not report sexual abuse more generally. Top 10 Reasons People Do Not Report Sexual Abuse. (2021, November 04). Retrieved November 17, 2021, from https://www.dlawgroup.com/reasons-people-do-not-report-sexual-abuse/

8 Chapter 3: Sex Offender Typologies. (n.d.). Retrieved November 17, 2021, from https://smart.ojp.gov/somapi/chapter-3-sex-offender-typologies

9 Finkelhor, David, The Prevention of Childhood Sexual Abuse, Retrieved November 17, 2021, from http://unh.edu/ccrc/pdf/CV192.pdf

10 Wortley, R., Leclerc, B., Reynald, D. M., & Smallbone, S. (2019). What Deters Child Sex Offenders? A Comparison Between Completed and Noncompleted Offenses. Journal of Interpersonal Violence, 34(20), 4303-4327. doi:10.1177/0886260519869235; Finkelhor, David, Juveniles Who Commit Sex Offenses Against Minors, <u>Juvenile Justice Bulletin</u>, December 2009, Retrieved November 17, 2021, from https://www.ojp.gov/pdffiles1/ojjdp/227763.pdf

11 Martin, E. K., & Silverstone, P. H. (2016). An Evidence-Based Education Program for Adults about Child Sexual Abuse (“Prevent It!”) That Significantly Improves Attitudes, Knowledge, and Behavior. *Frontiers in Psychology, 7*. doi:10.3389/fpsyg.2016.01177

12 Id. (References omitted)

13 Id. (References omitted)

14 Gubbels, J., van der Put, C.E., Stams, GJ.J.M. et al. Effective Components of School-Based Prevention Programs for Child Abuse: A Meta-Analytic Review. Clin Child Fam Psychol Rev 24, 553–578 (2021). https://doi.org/10.1007/s10567-021-00353-5

15 Stoltenborgh, M., Bakermans-Kranenburg, M. J., Alink, L. R. A., & Van IJzendoorn, M. H. (2015). The prevalence of child maltreatment across the globe: Review of a series of meta-analyses. Child Abuse Review, 24, 37–50. https://doi.org/10.1002/car.2353

16 Martin, E. K., & Silverstone, P. H. (2016). An Evidence-Based Education Program for Adults about Child Sexual Abuse (“Prevent It!”) That Significantly Improves Attitudes, Knowledge, and Behavior. *Frontiers in Psychology, 7*. doi:10.3389/fpsyg.2016.01177

17 But see Gibson, L. E., & Leitenberg, H. (2000). Child sexual abuse prevention programs: Do they decrease the occurrence of child sexual abuse? Child Abuse and Neglect, 24(9), 1115–1125. https://doi.org/10.1016/s0145-2134(00)00179-4; Diaz, M. J., Wolfersteig, W., Moreland, D., Yoder, G., Dustman, P., & Harthun, M. L. (2020). Teaching Youth to Resist Abuse: Evaluation of a Strengths-Based Child Maltreatment Curriculum for High School Students. *Journal of Child & Adolescent Trauma*, 14(1), 141-149. doi:10.1007/s40653-020-00304-2; Pulido, M. L., Dauber, S., Tully, B. A., Hamilton, P., Smith, M. J., & Freeman, K. (2015). Knowledge Gains Following a Child Sexual Abuse Prevention Program Among Urban Students: A Cluster-Randomized Evaluation. *American Journal of Public Health*, 105(7), 1344-1350. doi:10.2105/ajph.2015.302594; Collin-Vézina, D., Daigneault, I., & Hébert, M. (2013). Lessons learned from child sexual abuse research: Prevalence, outcomes, and preventive strategies. *Child and Adolescent Psychiatry and Mental Health*, 7(1), 22. doi:10.1186/1753-2000-7-22; Mikton, C., & Butchart, A. (2009). Child maltreatment prevention: A systematic review of reviews. *Bulletin of the World Health Organization*, 87(5), 353-361. doi:10.2471/blt.08.057075; Reynolds, A. J., Mathieson, L. C., & Topitzes, J. W. (2009). Do Early Childhood Interventions Prevent Child Maltreatment? *Child Maltreatment*, 14(2), 182-206. doi:10.1177/1077559508326223; Walsh, K., Zwi, K., Woolfenden, S., & Shlonsky, A. (2015). School-Based Education Programs for the Prevention of Child Sexual Abuse. *Research on Social Work Practice*, 28(1), 33-55. doi:10.1177/1049731515619705 (updated in 2018)

18 Martin (2016)

19 Risk Factors. (n.d.). Retrieved August 16, 2021, from https://www.inspq.qc.ca/en/sexual-assault/understanding-sexual-assault/risk-factors

20 Vallett, J. D. (2019). The Diffusion of Erins Law: Examining the Role of the Policy Entrepreneur. Academy of Management Proceedings, 2019(1), 10091. doi:10.5465/ambpp.2019.10091abstract

21 The State of America’s Children 2021 - Child Population. (2021, March 28). Retrieved August 22, 2021, from https://www.childrensdefense.org/state-of-americas-children/soac-2021-child-population/

22 NCANDS. (n.d.). Retrieved August 16, 2021, from https://www.acf.hhs.gov/cb/research-data-technology/reporting-systems/ncands

23 https://www.enoughabuse.org/images/Legislation/A_Call_to_Action-June2021.pdf

24 NCANDS Child File Code Book - NDACAN. (2021, July 23). Retrieved November 19, 2021, from https://ndacan.acf.hhs.gov/datasets/pdfs_user_guides/ncands-child-file-codebook.pdf

25 Cunningham, S. (2021). Causal Inference: The Mixtape. NEW HAVEN; LONDON: Yale University Press. doi:10.2307/j.ctv1c29t27

26 Goodman-Bacon, A. (2018). Difference-in-Differences with Variation in Treatment Timing. National Bureau of Economic Research. doi:10.3386/w25018

27 Callaway, B., & Sant’Anna, P. H. (2020). Difference-in-Differences with multiple time periods. Journal of Econometrics. doi:10.1016/j.jeconom.2020.12.001

28 Their methodology is described here: https://bcallaway11.github.io/did/articles/multi-period-did.html. Sample code in r using the so called “did” package can be found here: https://bcallaway11.github.io/did/articles/did-basics.html

29 Borusyak, K., Jaravel, X., & Spiess, J. (2021, August 27). Revisiting Event Study Designs: Robust and Efficient Estimation. Retrieved December 1, 2021, from https://arxiv.org/abs/2108.12419

30 Jacob, Daniel, CATE meets ML - Conditional Average Treatment Effect and Machine Learning (March 30, 2021). Available at SSRN: https://ssrn.com/abstract=3816558 or http://dx.doi.org/10.2139/ssrn.3816558

31 Naushan, Haaya, Causal Machine Learning for Econometrics: Causal Forests, TowardDataScience, April 2021, Retrieved November 17, 2021, from https://towardsdatascience.com/causal-machine-learning-for-econometrics-causal-forests-5ab3aec825a7?gi=363aa27a981d

32 McKenzie, D. (2020, January 21). Revisiting the Difference-in-Differences Parallel Trends Assumption: Part I Pre-Trend Testing [Web log post]. Retrieved November 17, 2021, from https://blogs.worldbank.org/impactevaluations/revisiting-difference-differences-parallel-trends-assumption-part-i-pre-trend

33 Introduction to DiD with Multiple Time Periods. (n.d.). Retrieved November 17, 2021, from https://bcallaway11.github.io/did/articles/multi-period-did.html

34 Marcus, M., & Sant’Anna, P. H. (2021, March 01). The Role of Parallel Trends in Event Study Settings: An Application to Environmental Economics: Journal of the Association of Environmental and Resource Economists: Vol 8, No 2. Retrieved November 17, 2021, from https://www.journals.uchicago.edu/doi/10.1086/711509

35 Chapter 2, Child Maltreatment 2019, Report by the United States Department of Health and Human Services. January 14, 2021 accessed December 17, 2021 at https://www.acf.hhs.gov/cb/report/child-maltreatment-2019

36 Research Paper of the Department of Health and Human Services, Decision-Making in Unsubstantiated Child Protective Services Cases: Synthesis of recent research, June 2003, at https://www.childwelfare.gov/pubpdfs/decisionmaking.pdf

37 Note that throughout this paper we are only talking about the formal, legal reporting mechanism for child abuse including CSA. There is no question that a child who is experiencing grooming should report that fact to a responsible adult. However, as grooming is not illegal by itself, it should not be reported through the official child abuse reporting mechanisms because it is not child abuse. It may be the case that some states do not have a clear pathway for reporting grooming and other such behaviors that do not in and of themselves constitute CSA. To avoid all of the potential repercussions of reporting activity as CSA when it is not, states should adopt a mechanism specific to these activities that are not illegal but nonetheless deserve some sort of monitoring.

38 Children’s Bureau, United States Department of Health and Human Services, Disclosure of Confidential Child Abuse and Neglect Records: STATE STATUTES, current through June 2017, accessed at https://www.childwelfare.gov/pubpdfs/confide.pdf on December 29, 2021.

39 https://andrewcbaker.netlify.app/2019/09/25/difference-in-differences-methodology/

